# Bivariate GSA-MiXeR: A Novel Tool for Functional Genomic Analyses Implicates Diverse Neural Cell Types for Psychiatric and Neurodegenerative Disorders

**DOI:** 10.64898/2025.12.16.25342384

**Authors:** Nadine Parker, Julian Furher, Dat Nguyen, Vera Fominykh, Piotr Jaholkowski, Ibrahim Akkouh, Kevin S. O’Connell, Espen Hagen, Shahram Bahrahmi, Dag Årsland, Sverre Bergh, Torgeir Engstad, Tormod Fladby, Anne-Brita Knapskog, Karin Persson, Gøril Rolfseng Grøntvedt, Bengt-Ove Madsen, Arvid Rongve, Ingvild Saltvedt, Sigrid B. Sando, Katja Scheffler, Geir Selbæk, Eystein Stordal, Mathias Toft, Leiv Otto Watne, Srdjan Djurovic, Olav B. Smeland, Anders M. Dale, Alexey A. Shadrin, Ole A. Andreassen, Oleksandr Frei

## Abstract

The growing number of genomic discoveries in complex human traits has highlighted the need for advanced functional genomics tools that parse their polygenic and pleiotropic genetic architecture to provide biological insights. We present bivariate GSA-MiXeR, a novel tool that models the partitioned heritability and covariance of two traits within a genomic region of interest (ROI) and estimates the (i) trait-specific fold enrichment, (ii) local genetic correlation, and (iii) local genetic omnibus statistic for ranking genomic ROIs. Unlike previous methods, our tool estimates local genetic correlations both in continuous and disjoint genomic ROIs, expanding the ability to assess local genetic overlap among complex traits. We perform simulations to validate our tool and illustrate its utility in applied analyses that implicate diverse neural cell types for psychiatric and neurodegenerative disorders using single cell RNA sequencing data. Bivariate GSA-MiXeR provides new analytical avenues that facilitate a transition from genetic discovery to mechanistic insights.

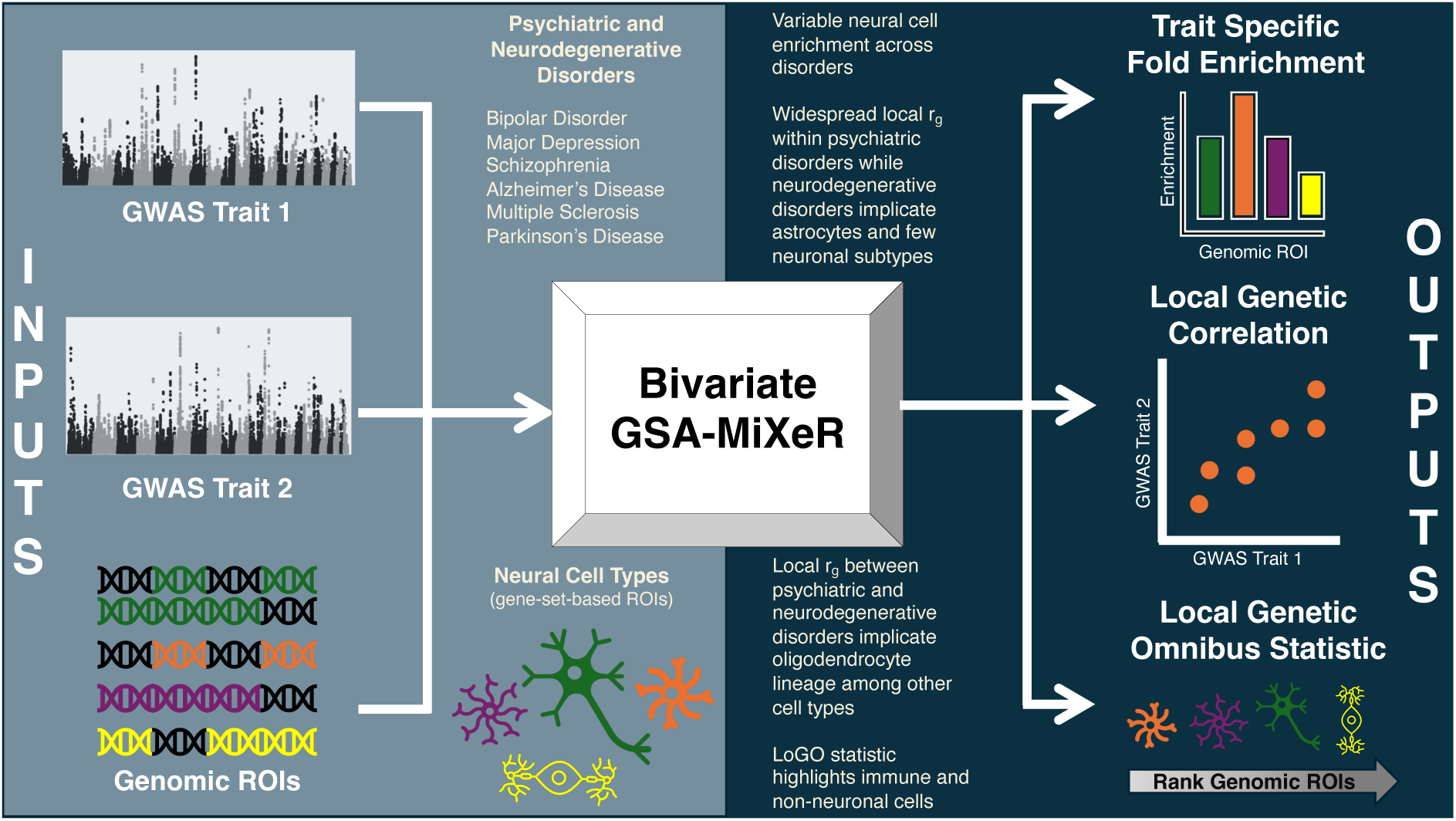

## Introduction

Two-decades of genome-wide association studies (GWAS) have revealed that many human traits are polygenic; with numerous associated genetic variants, and from this pervasive polygenicity follows pleiotropy; where many genetic variants are strongly associated with multiple complex traits.^1^ The functional genomic characterization of polygenic traits and their pleiotropic genetic effects is crucial for translating GWAS findings to relevant mechanisms. The parsing of polygenicity to provide biological insights is a common approach with some methods partitioning heritability across functionally relevant genomic regions to estimate enrichment in GWAS traits.^2,3^ In line with this approach, we previously developed the gene set analysis MiXeR (GSA-MiXeR) which implicates biologically specific gene sets and provides an estimate of heritability fold-enrichment.^3^ Genetic correlations (r_g_s) are perhaps the most common approach to assess genetic overlap, which can be indicative of local pleiotropic effects.^4^ Genome-wide r_g_s are frequently used in genomic studies of complex traits but this aggregate level statistic can mask more nuanced genetic overlap within localized regions of the genome.^4–6^ Recent methodological advances have allowed for a more granular dissection of r_g_ within localized regions that are likely driven by pleiotropic effects.^5,6^ However, these tools were originally designed to estimate local r_g_s in continuous regions of the genome and require low linkage disequilibrium (LD) between adjacent regions. In general, there is a need for methods which can parse both the polygenicity and extensive genetic overlap of complex traits to further facilitate the transition from locus discovery to functional biological insights.

Here we present bivariate GSA-MiXeR, a novel tool which extends GSA-MiXeR^3^ to model two traits simultaneously. Bivariate GSA-MiXeR uses GWAS summary statistics and pre-defined genomic regions of interest (ROIs; e.g., gene sets) as input and provides (i) trait-specific fold enrichments, (ii) a local r_g_, and (iii) a local genetic omnibus (LoGO) statistic, for each genomic ROI as output. The novel bivariate estimates enable a broader functional assessment of pairs of complex polygenic traits and their pleiotropic effects. In contrast to other methods, bivariate GSA-MiXeR expands the utility of local r_g_ estimation beyond continuous LD-based ROIs to additionally encompass disjoint regions of the genome, including gene sets with genes distributed across chromosomes. In addition, the LoGO statistic allows for the ranking of the genomic ROIs based on their importance to both traits. Therefore, our novel bivariate GSA-MiXeR tool offers unique opportunities for functional genomic characterization of pairs of traits.

In applied analyses, we use bivariate GSA-MiXeR to investigate the shared and distinct neural cell type associations among psychiatric and neurodegenerative disorders. These disorders are polygenic, exhibit genetic overlap, and comorbidity is observed between the two clinical domains.^7–9^ Molecular profiling of these disorders consistently implicate brain tissue and neurobiological processes.^3,8,10–15^ The surge in single-cell and single-nucleus RNA sequencing (scRNA-seq and snRNA-seq) studies of the human brain has facilitated more granular investigations of these brain-related disorders. However, to date, these studies have primarily been performed through univariate analyses. Functional genomic analyses using GWAS of common psychiatric disorders, such as bipolar disorder (BD), major depression (MD), and schizophrenia (SZ) often implicate various subtypes of neurons.^8,10,14,16–19^ Meanwhile, GWAS of disorders which feature neurodegeneration (neurodegenerative disorders), such as Alzheimer’s disease (AD), multiple sclerosis (MS) and Parkinson’s disease (PD) tend to implicate immune cells like microglia and a smaller subset of neuronal subtypes.^8,13,16,17,20,21^ These cell-specific distinctions help reveal biological features that elucidate how conditions implicating the same organ manifest in differing clinical presentation and may inform further clinical subtyping and treatment. By applying bivariate GSA-MiXeR we can jointly examine these disorders with cell type specificity, allowing us to understand their shared and distinct mechanisms and elucidate putative biological underpinnings of comorbidity.

## Results

### Summary of Bivariate GSA-MiXeR and Overview of Analyses

The main inputs to bivariate GSA-MiXeR includes two sets of GWAS summary statistics and a pre-defined list of genomic ROIs. Direct optimization of the bivariate GSA-MiXeR likelihood function is used to estimate model parameters leveraging GWAS z-scores for single nucleotide polymorphisms (SNPs) (see Methods). For each GWAS trait and a given genomic ROI, an estimated fold enrichment of heritability is produced, defined as the proportion of the trait’s SNP heritability in the genomic ROI divided by the expected proportion of SNP heritability under a baseline model agnostic to that genomic ROI. Also, within a given genomic ROI, a local r_g_ is estimated along with a p-value (for r_g_ being different from zero) derived from a likelihood ratio test (see Methods). Additionally, a LoGO statistic is generated which is the log-likelihood difference between a bivariate model that incorporates ROI-specific parameters for both traits and a model that is agnostic to the ROI. Therefore, the LoGO statistic serves as an omnibus test for whether the genomic ROI (i) is enriched for one or both traits and/or (ii) has a local r_g_ between the two traits. The univariate partitioning of heritability and bivariate partitioning of covariance serve separate functionality. Therefore, a significant local r_g_ does not require the presence of enrichment in either trait. However, both the univariate (fold enrichment) and bivariate (r_g_ and LoGO statistic) metrics can be used for functional characterization of the input GWAS traits.

We performed a series of simulations to validate bivariate GSA-MiXeR estimates of local genetic correlation and conducted a method comparison. These simulations used genomic ROIs that represent (i) continuous LD-based regions that allow for comparison with existing methods and (ii) disjoint regions using randomly generated gene sets. In applied analyses, we used GWAS for pairs of psychiatric (i.e., BD, MD, SZ) and neurodegenerative (i.e., AD, MS, PD) disorders with genomic ROIs defined by gene sets for diverse neural-cell-types.^10,14,18,15,20,13,22–24^ To translate our findings to inform individual-level liability to these disorders, we assessed the association between bivariate GSA-MiXeR estimates and the performance of cell-type-based polygenic risk scores (PRSs). Finally, we further illustrated the utility of bivariate GSA-MiXeR to implicate more granular functional biology through analyses with single-cell expression quantitative trait loci (sc-eQTL) data.

### Bivariate GSA-MiXeR Accurately Estimates Local Genetic Correlations

We conducted a series of simulations to assess the ability of bivariate GSA-MiXeR to recover the true local r_g_ between two traits (i.e., correlation in known causal effect sizes). The first set of simulations was designed to (i) benchmark bivariate GSA-MiXeR estimation of local r_g_ against another method and (ii) assess the accuracy of bivariate GSA-MiXeR local estimation of r_g_ using randomly generated gene sets comprised of disjoint regions of the genome. For these analyses we used simulated pairs of continuous traits with equal heritability (ℎ^2^ = 0.3) and polygenicity (𝜋 = 0.003 ) and with 50% overlap in causal SNPs. We additionally vary the correlation in effect sizes of shared causal SNPs from −1 to 1 using 0.25 increments using a random effects model.

We compared the performance of bivariate GSA-MiXeR to the local version of the high-definition likelihood (HDL-L) tool^6^ based on the estimation of local r_g_. Although other methods for the estimation of local r_g_ exist,^5,25,26^ both bivariate GSA-MiXeR and HDL-L use a random effects model with parameters estimated using a likelihood function. Therefore, the two methods have a comparable approach to estimation of local r_g_. Preexisting tools were originally designed to estimate local r_g_ within continuous genomic ROIs typically defined based on LD structure. Therefore, here we use the 34 approximately independent LD-based genomic regions on chromosome 22, originally designed for HDL-L use, as our predefined genomic ROIs. Simulated pairs of traits (n=1,530) were designed to have 20% of their shared causal SNPs selected from a given genomic ROI. Bivariate GSA-MiXeR and HDL-L had comparable performance in estimating the local r_g_ within the 34 continuous genomic ROIs (Figure 1A-C). To estimate the variability in accuracy between methods, we calculated the average mean squared error (MSE) within each of the 34 genomic ROIs and found no significant difference between the two methods across those ROIs (t(33)=0.78, p=0.44, Supplementary Table 1) and the average MSE across all simulation runs was comparable (Mean MSE_bivariateGSA- MiXeR_=0.04; Mean MSE_HDL-L_=0.03). Across all simulations, the correlation in estimated vs true local r_g_ was high for both methods (Figure 1B; bivariate GSA-MiXeR r_g_=0.95, p<0.001 ; HDL- L r_g_=0.96, p<0.001) and strongly correlated between the two methods (Figure 1C; r=0.96, p<0.001).

**Figure 1.**
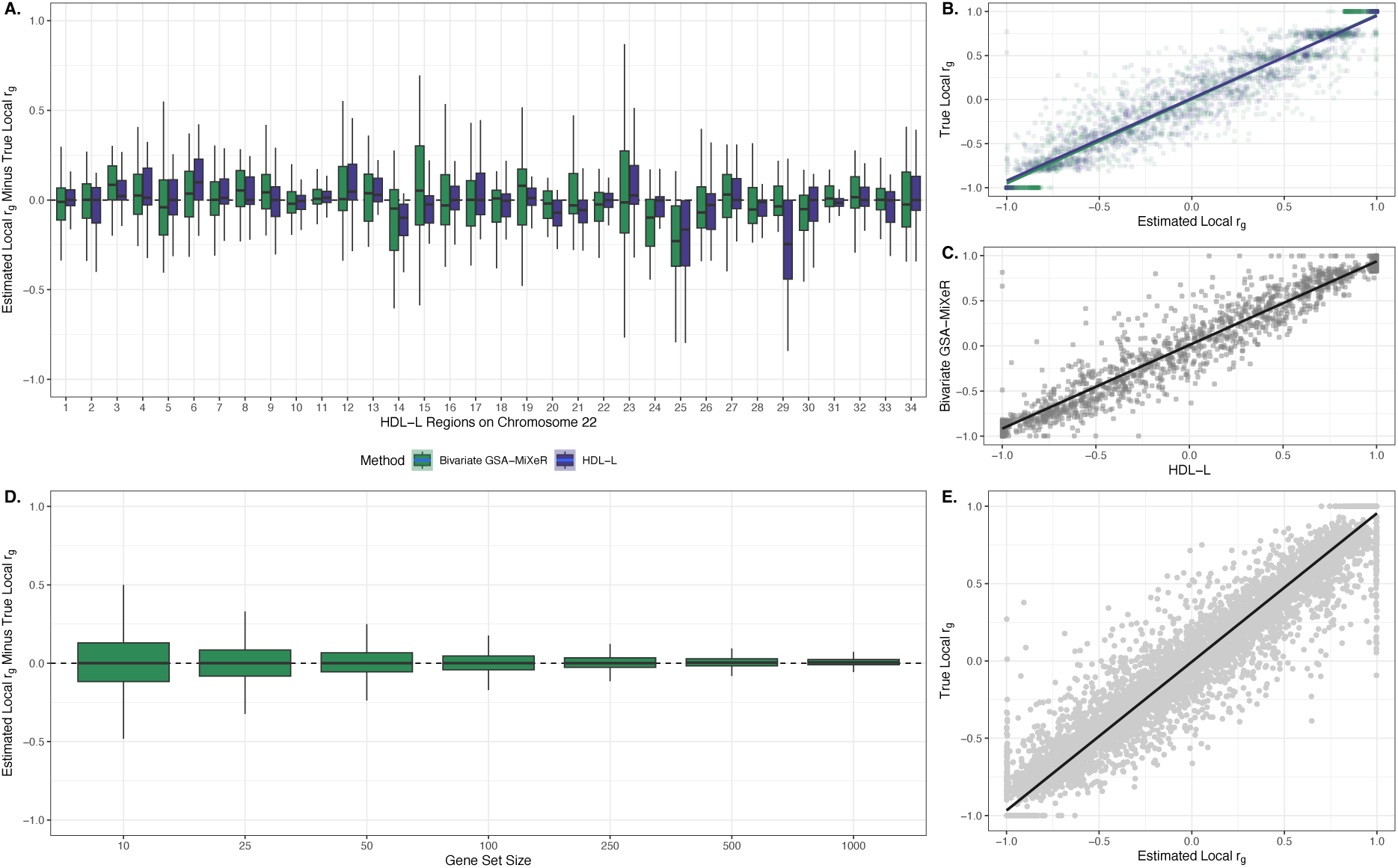
**Simulation results**. A) The box plots show the difference in estimated and true local genetic correlation within 34 continuous genomic ROIs on chromosome 22 for bivariate GSA-MiXeR (green) and HDL-L (blue) for all simulated scenarios. B) A scatter plot depicting the association between the true local genetic correlation in simulated betas and the estimated local genetic correlation by bivariate GSA-MiXeR (green) and HDL-L (blue) across all simulations. C) The comparison of genetic correlation estimated by bivariate GSA-MiXeR and HDL-L across all simulations. D) Box plots showing the difference in the true local genetic correlation and that estimated by bivariate GSA-MiXeR within gene sets of varying size. E) The association between the true local genetic correlation and that estimated by bivariate GSA-MiXeR across all simulations using gene sets as genomic regions.

In simulations of disjoint regions of the genome, we used genes that contained at least one causal SNP shared between simulated traits to generate random gene sets of varying size (n=10, 25, 50, 100, 250, 500, and 1000 genes). These simulations reveal strong alignment between the true local r_g_ values and bivariate GSA-MiXeR estimates (Figure 1D-E, r=0.98, p<0.001). In addition, these findings illustrate the novel utility of our method to estimate local r_g_ unrestricted to continuous regions of the genome.

In a separate set of simulations, we assessed the accuracy of bivariate GSA-MiXeR under null simulations. Here, we simulated two traits with 100% overlap in causal SNPs, an r_g_=0, and heritability localized to a single gene set of 100 randomly selected genes where heritability was informed by previous real data analysis (ℎ^2^ = 10^−4^ to 10^−3^).^3^ We repeat this simulation procedure 1000 times at each heritability level. We find bivariate GSA-MiXeR p-value estimates are well calibrated under null (Supplementary Figure 1).

### Variable Cell Type Enrichment Exhibited Across Brain-Related Disorders

We applied bivariate GSA-MiXeR to GWAS of psychiatric and neurodegenerative disorders using genomic ROIs defined by cell-type gene sets. To define cell-type gene sets we used data from an advanced large-scale census of the cellular transcriptome derived from either (i) the early prenatal brain using snRNA-seq,^22^ (ii) the adult brain^24^ or (iii) adult cortex^23^ using scRNA-seq. For the early prenatal and adult brain data sources, we used the highest order classification of cell type defined by the original studies (n_adult_= 31 superclusters and n_prenatal_=12 classes) and generated cell type gene sets using genes with a high expression proportion for that cell type (see Methods for details). For the adult cortex sample, we selected marker genes conserved across eight sampled cortical regions for 24 subclasses of cells, in line with a previous approach.^27^ Therefore, the adult brain source provides brain-wide data with regional specificity of the cellular transcriptome but, the adult cortex source provides consistent signatures of the cellular transcriptome across the cortex implicating diffuse cortical involvement for a given cell type. Overall, our applied analyses contrast two clinical domains of brain-related disorders and neural cell types with differing temporal and spatial contexts.

Across the 67 cell type gene sets, there was a wide variation in both fold enrichment and depletion of heritability for the six disorders (Figure 2A, Supplementary Table 2). Each psychiatric disorder was most strongly enriched for a specific cortical inhibitory neuron: Pax6 expressing interneurons for BD (fold enrichment=1.58, se=0.25), somatostatin (SST) expressing interneurons for MD (fold enrichment=1.75, se=0.14), and parvalbumin (Pvalb) expressing interneurons for SZ (fold enrichment=1.89, se=0.25). These findings are supported by previous studies implicating cortical neurons that express these marker genes using induced pluripotent stem cells, in situ hybridization with post-mortem human cortical brain tissue, and alternative GWAS-based enrichment approaches.^16,28–31^ The three psychiatric disorders clustered together based on their pattern of fold enrichment, generally more enriched for neuronal cell types and depleted for other cell types (Figure 2B, Supplementary Table 3). In line with previous reports,^16^ AD was enriched for several immune related cell types and along with MS had the strongest fold enrichment for the immune class of cells sampled from early prenatal brain tissue (AD: fold enrichment=2.36, se=0.20; MS: fold enrichment=1.79, se=0.12). PD had strong enrichment for a mixture of neuronal and non-neuronal cell types with the strongest enrichment observed for neurons sampled from the early prenatal brain (fold enrichment=2.59, se=0.30). The strong PD neuronal enrichment and differentiation in the pattern of enrichment from AD is in line with previous reports.^8,13,32^ Notably, PD exhibited the largest average enrichment for cell types derived from early prenatal brain tissue across the six disorders (Supplementary Table 4). Overall, these univariate fold enrichment findings show general consistency with previous reports providing validation through replication.

**Figure 2.**
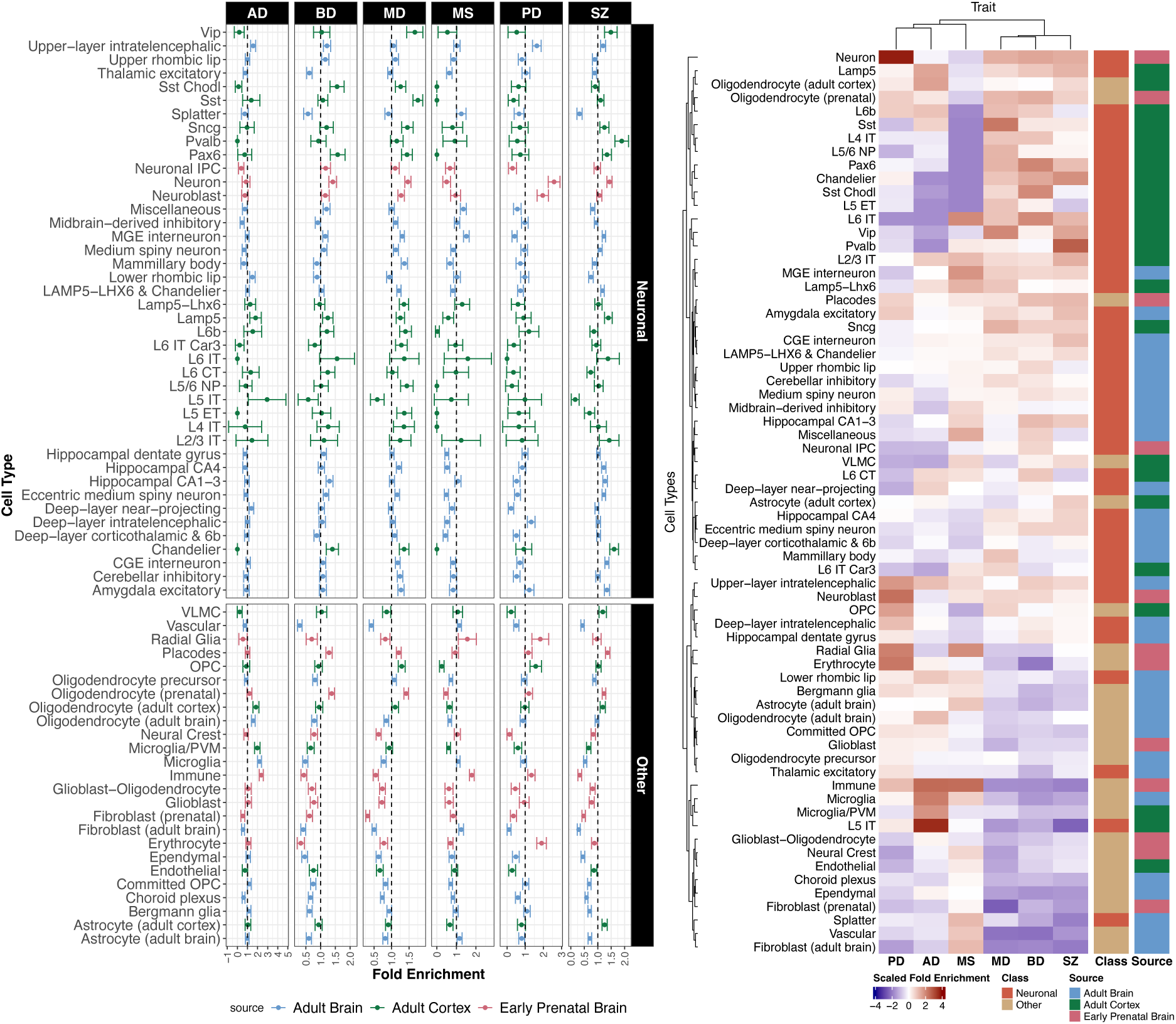
**Disorder fold enrichment for neural cell types**. A) The fold enrichment and standard errors are depicted for each cell type across the six brain-related disorders. The dotted line references a fold enrichment of one. B) Hierarchical clustering of the scaled fold enrichment. Prior to clustering, fold enrichment values were mean centered and scaled to a have a standard deviation of one across all cell types for a given disorder.

### Local Genetic Correlations Implicate Novel Transdiagnostic Cell Type Associations

Genome-wide r_g_ between pairs of psychiatric and neurodegenerative disorders as well as local r_g_ within continuous LD-defined ROIs have previously been reported.^8,33^ Here, we performed novel analyses assessing local r_g_ within cell type gene sets representing disjoint regions of the genome of functional relevance. In doing so, we observed pervasive local genetic overlap between pairs of psychiatric disorders across cell types (Figure 3, Supplementary Table 5). For BD vs SZ and BD vs MD 66 of the 67 cell types tested had a local r_g_ that survived correction for multiple comparisons while MD vs SZ had 63. The strength of the local r_g_ within gene sets hovered around the genome-wide r_g_ (blue line in Figure 3). However, the strongest correlations for BD vs SZ as well as BD vs MD was in the L2/3 intratelencephalic (IT) cell type (BD vs SZ: r_g_ =0.94, p_fdr_=6.33e-4; BD vs MD: r_g_ =1.00, p_fdr_=1.43e-5) and for MD vs SZ was in the vascular leptomeningeal cells (VLMC; r_g_ =0.72, p_fdr_=3.87e-16) gene set which deviated most from the genome-wide r_g_. IT neurons can form corticocortical and corticostriatal projections important for motivated behaviors and have been linked to many brain-related disorders.^32,34,35^ Meanwhile, the strong VLMC correlation between MD and SZ may suggest a shared role for vascular disruptions to the blood brain barrier in disorder pathology.^36–39^ For the neurodegenerative disorders, only AD vs MS and AD vs PD had local r_g_s that survived correction for multiple comparisons within four cell types (Figure 3B). Again, upper-layer IT neurons were implicated for AD vs PD (r_g_ =0.33, p_fdr_=0.03). Additionally, local r_g_s between AD and MS as well as AD and PD were observed within astrocyte gene sets (including Bergmann glia) sampled from adult brain or cortex. This highlights the putative shared role of astrocytes with previous suggestions of their pro-inflammatory mechanisms, altered metabolism, and disrupted molecular homeostasis contributing to the progression of neurodegeneration.^40,41^

**Figure 3:**
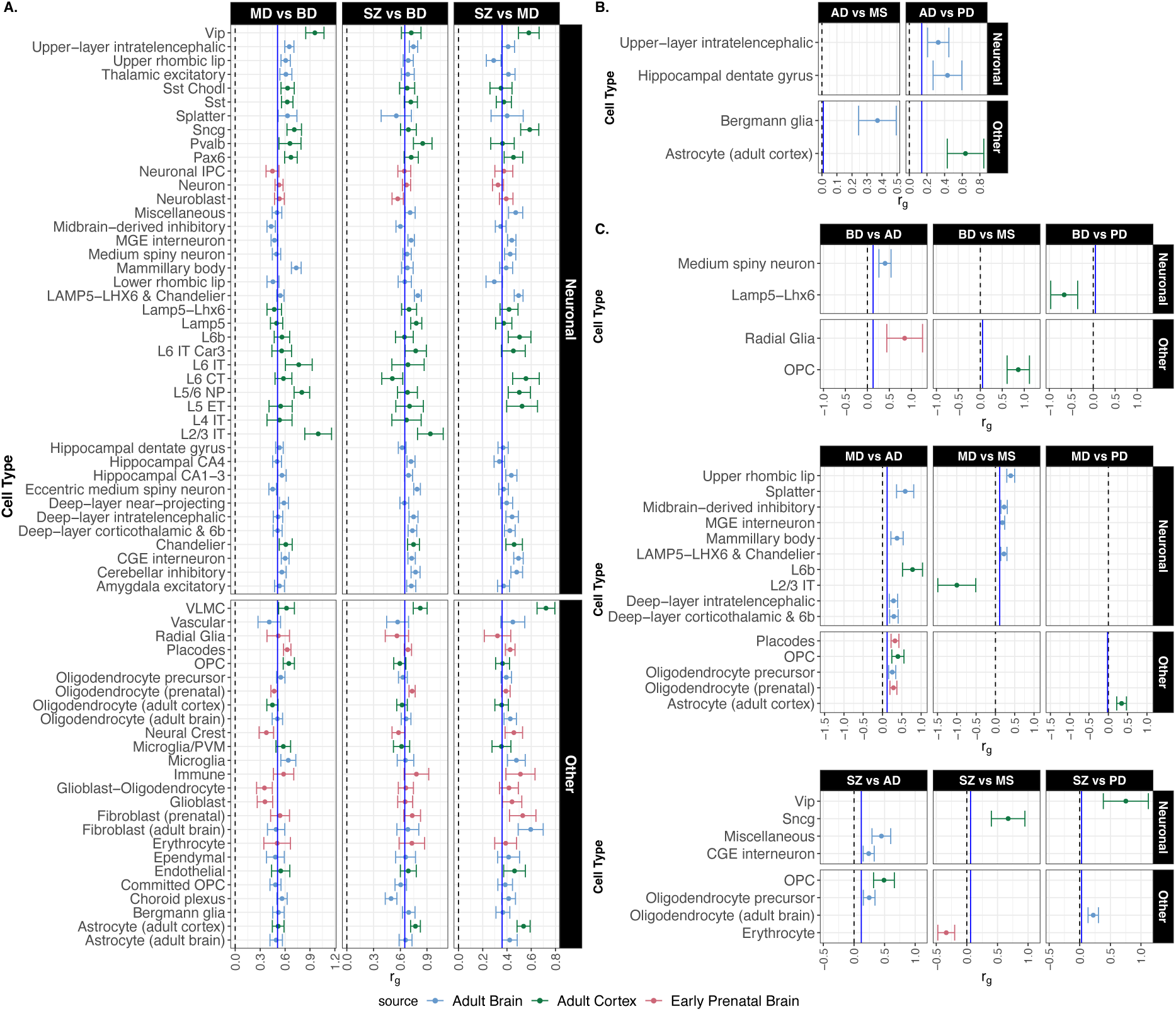
**Cell type local genetic correlations across pairs of disorders**. Local genetic correlations between pairs of A) psychiatric disorders, B) neurological disorders, and C) psychiatric and neurological disorders within the cell type gene sets. Error bars are equivalent to plus or minus one standard error. Note, only significant genetic correlations after correction for multiple comparisons are presented (i.e., false discovery rate q-value < 0.05). The absence of a point estimate means the correlation did not meet this threshold for significance. The solid blue line represents the genome-wide genetic correlation between the pairs of disorders. The dotted black line references a genetic correlation of zero.

Among the local r_g_s between psychiatric and neurological disorders, MD vs AD had the most associations (n=9 cell types) which may reflect their common comorbidity and the advanced neuropathology observed in comorbid cases.^42,43^ For MD vs AD, the strongest r_g_ was for L6b cortical neurons (r_g_=0.78, p_fdr_=1.20e-3). Neurons in this deepest layer of the cortex form corticocortical and corticothalamic connections involved in modulating brain state.^44^ Of note, oligodendrocyte-related cell types were implicated in 7 of the significant local r_g_s between psychiatric and neurological disorders encompassing 63.63% of the significant non-neuronal associations. The widespread implication of the oligodendrocyte lineage may reflect the shared importance of effective neurotransmission and interregional brain connectivity, processes supported by myelination, in the pathophysiology of these disorders.^12,45–47^ Unlike the solely positive local r_g_s observed between disorders in the same clinical domains, there were three negative local r_g_s observed between pairs of psychiatric and neurodegenerative disorders. Here, the L2/3 IT cell type is again implicated this time with a strong negative r_g_ between MD and MS (r_g_=-1.00, p_fdr_=0.02). Also, for BD and PD as well as SZ and MS a negative local r_g_ was observed in Lamp-Lhx6 neuronal cells (r_g_=-0.66, p_fdr_=0.04) and erythrocytes (r_g_=-0.34, p_fdr_=4.68e-2), respectively. These negative local r_g_s suggest the same cell type may have opposing effects in each of the paired disorders. We also clustered z-scores for the pair-wise local r_g_ analyses (accounting for strength, direction, and uncertainty in estimates). The resulting clusters were predominantly driven by the local r_g_ between pairs of psychiatric disorders (Figure 4A).

**Figure 4:**
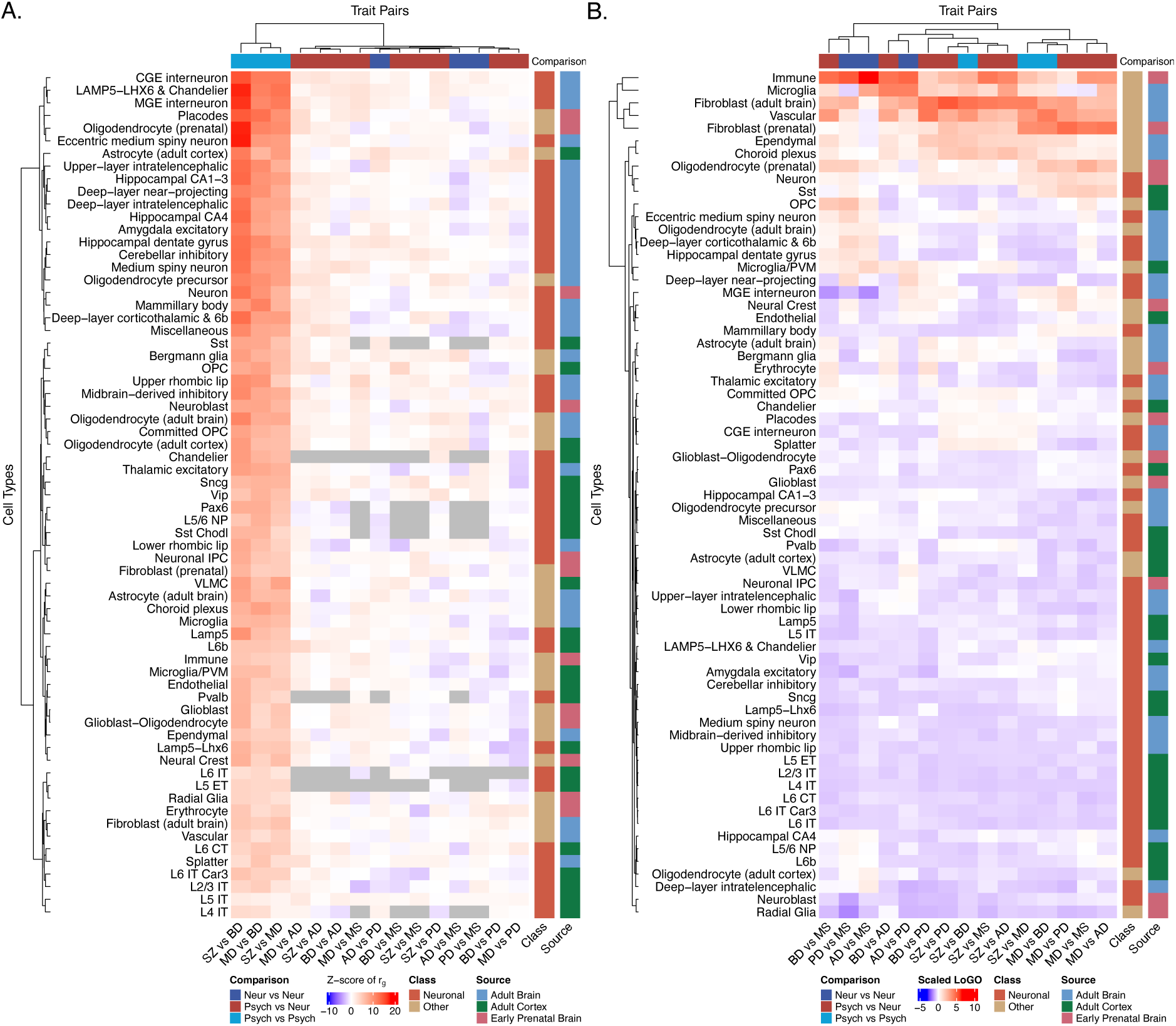
**Clustering local genetic correlations and omnibus statistics**. A) Hierarchical clustering of the z-score for the local genetic correlations within cell type gene sets. Grey represents genetic correlations that were not estimated due to lack of genetic signal. B) Hierarchical clustering of the scaled LoGO statistic (mean centered and standard deviation of one) within cell type gene sets.

Perhaps the strongest evidence for a given cell type having a shared role in the pathophysiology of two disorders is when there is an observed local r_g_ and both traits exhibit fold enrichment for that cell type. This scenario may be indicative of pleotropic effects within the genomic ROI and occurs frequently among pairs of psychiatric disorders. BD, MD, and SZ were all enriched and pairwise correlated among: Chandelier, Eccentric medium spiny neuron, Lamp5, medial ganglionic eminence (MGE) interneurons, Pax6 neurons from adult brain/cortex gene sets as well as neurons, placodes, and oligodendrocytes from early prenatal brain gene sets. The scenario also occurs once for pairs of neurological traits with AD vs PD in upper-layer IT neurons. However, this scenario of putative pleiotropy was not observed for the psychiatric and neurodegenerative disorder pairings for any cell type.

### The LoGO Statistic Highlights Bivariate Enrichment/Depletion for Non-neuronal Cells

We used the LoGO statistic, a metric which reflects the relative importance of a genomic ROI for a pair of traits, to rank the cell-type gene sets for pairs of disorders. This ranking based on the LoGO statistic was primarily driven by strong enrichment or depletion for a given pair of disorders (Supplementary Figure 2-16). For pairs of psychiatric disorders, the LoGO statistic was highest in cell types which exhibited strong depletion in both disorders and a significant local r_g_, such as fibroblasts (LoGO_SZvsMD_=173.88, LoGO_SZvsBD_=160.10) and vascular (LoGO_BDvsMD_=131.97) cells. These LoGO statistic rankings suggested the strength of depleted genetic signal in these cell types outweighs any shared enriched genetic signal among the pairs of psychiatric disorders. In contrast, for neurodegenerative disorders, the LoGO statistic was highest in cell types which exhibited enrichment for both disorders, such as the early prenatal immune cell type (LoGO_ADvsMS_=107.52, LoGO_ADvsPD_=56.62, LoGO_MSvsPD_=58.88), highlighting the neuroimmune-related biology in neurodegenerative pathology.^48–51^ For pairs of psychiatric and neurodegenerative disorders, immune related, vascular, and subsets of neurons were highly ranked based on combinations of strong enrichment and depletion of genetic signal (Figure 3E, Supplementary Figure 8-16). When performing hierarchical clustering based on the LoGO statistic, patterns appear based on gene sets (Figure 4B) with non-neuronal cell types highly ranked across the pairs of disorders. This pattern of clustering based on LoGO statistic, contrasts that observed when clustering based on local r_g_ (Figure 4A) highlighting the separate utility of the two bivariate statistics (local r_g_ vs LoGO statistic) for understanding local genetic architecture among complex traits.

### Bivariate GSA-MiXeR Metrics Relate to Polygenic Risk Score Performance

Revealing the partitioned heritability and covariance at a cell type level using bivariate GSA-MiXeR may be useful for a more granular understanding of individual-level liability to disorders. Therefore, we used a pathway polygenic risk score (PRS) approach and assessed the correlation between PRS performance in differentiating cases from controls and bivariate GSA-MiXeR metrics (see Methods for details). Briefly, we selected a subset of SNPs mapped to genes within the cell type gene set and used SNP effects estimated by MiXeR-Pred^52^ to generate cell type PRSs with PRSet^53^. Notably, MiXeR-Pred can produce standard univariate SNP effects or bivariate SNP effects (i.e., using a secondary trait to inform the effect estimate for the primary trait) for PRS calculations. We generated cell-type-specific PRS in an independent sample of 2863 AD cases, 620 PD cases, and 1791 controls for neurodegenerative disorders as well as a separate sample of 1070 BD cases, 1042 SZ cases, and 1422 controls for the psychiatric disorders.

At the univariate level, there were 40, 16, 48, 56, cell-type-specific PRS significantly associated with AD, PD, BD, SZ, respectively (Figure 5A, Supplementary Table 6, Supplementary Figure 17). For AD, immune, microglia, and astrocyte related PRSs explained a larger portion of phenotypic variance than a PRS which leveraged all SNPs (Figure 5A dotted line). This suggests there is reduced noise in differentiating cases and controls when restricting SNPs to genes highly expressed in these cell types. Also, the variance explained across the 40 associated AD cell type PRS was correlated with the AD fold enrichment estimates from bivariate GSA-MiXeR (r=0.36, p=0.02). However, this correlation was not observed for PD (r=0.07, p=0.80), BD (r=0.03, p=0.86), or SZ (r=0.25, p=0.07). These findings may reflect a strong genetically regulated role in disorder pathology for the top implicated cell types for AD, but less so for the other disorders.

**Figure 5.**
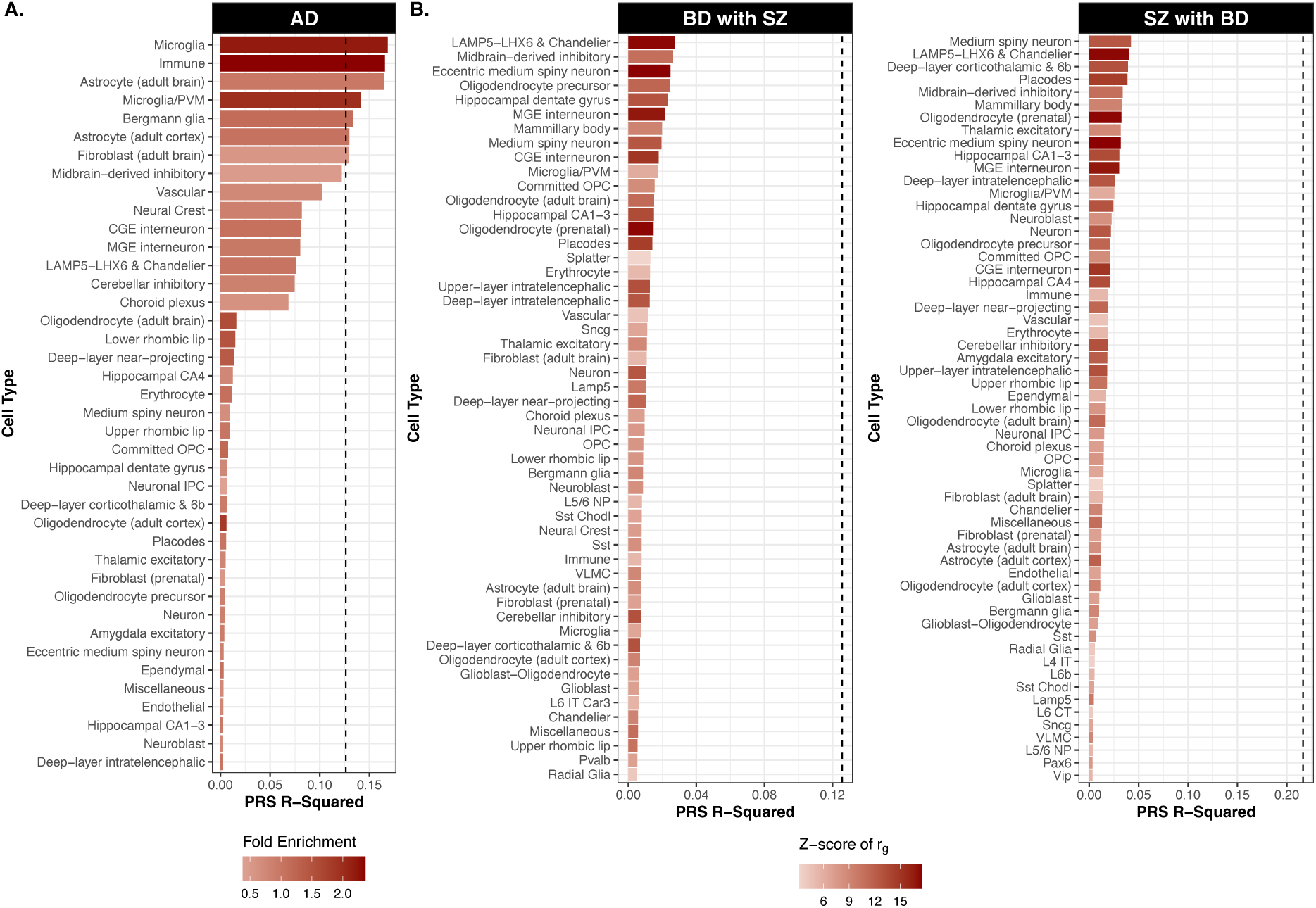
**Polygenic risk score performance and bivariate GSA-MiXeR metrics**. A) Variance explained (R-squared) by univariate Alzheimer’s disease (AD) cell type polygenic risk scores (PRS) with the intensity of the bar color represented by the strength of fold enrichment. B) Variance explained by bivariate, bipolar disorder (BD) with schizophrenia (SZ) and SZ with BD, cell type PRS where the secondary trait is selected based on the average absolute value of the z-score for local r_g_ across cell-type gene sets. The intensity of the bar color represents the strength of the local r_g_ between the two traits for the given cell type. The dotted line in each plot represents the variance explained for a PRS using all available SNPs.

At the bivariate level, we first selected a secondary trait based on the highest average absolute value of z-score for local r_g_ across all cell types (Supplementary Table 7) to generate PRS. Of those bivariate cell type PRS, 52 were significant associated with BD using SZ as a secondary trait and 59 with SZ using BD as a secondary trait (Figure 5B, Supplementary Table 8, Supplementary Figure 18). Here, the absolute value of z-scores for local r_g_ was correlated with PRS performance across cell types (BD with SZ: r=0.57, p=1.05e-05; SZ with BD: r=0.67, p=7.25e-09). This suggest that the extensive genetic overlap among psychiatric disorders can be used to inform prediction even with more localized cell type analyses which may be useful in defining subtypes which cross diagnostic boundaries. Though AD and PD had many associated bivariate cell type PRS, their prediction performance was not correlated with the absolute value of z-scores for local r_g_ (Supplementary Figure 18). In a separate bivariate analysis, we selected a secondary trait based on the highest average LoGO statistic (Supplementary Table 9). The performance of a bivariate cell type PRS for AD using SZ as a secondary trait correlated with the LoGO statistic (r=0.54, p=3.21e-4) but this association was not observed for the other disorders (Supplementary Table 10, Supplementary Figure 19). In general, the omnibus nature of the LoGO statistic may not be well suited for informing polygenic prediction.

### Bivariate GSA-MiXeR Links Genes to Disorders using Single Cell eQTL Data

To further illustrate the utility of bivariate GSA-MiXeR to implicate more granular functional biology, we estimated the local r_g_ between each of the six disorders and genes in a cell specific manner using data from cis-eQTL analyses of eight neural cell types^54^. Significant trait-gene correlations were observed for five cell types and four disorders after correction for multiple comparisons (Figure 6, Supplementary Table 11). MS had the most gene associations with predominantly negative correlations in genes expressed in oligodendrocyte progenitor cells and inhibitory neurons. The strongest local r_g_ was found between MD and AP5B1 expressed in pericytes. Pericytes and their role in regulating blood-brain barrier integrity have been linked to stress and inflammation in MD and other psychiatric disorders.^55,56^ Moreover, in a recent study of brain derived sc-eQTLs, Mendelian randomization analyses identified a similar positive association between AD and oligodendrocyte expression of complement receptor-1 (*CR1*), a gene with immune regulatory activity.^57^

**Figure 6.**
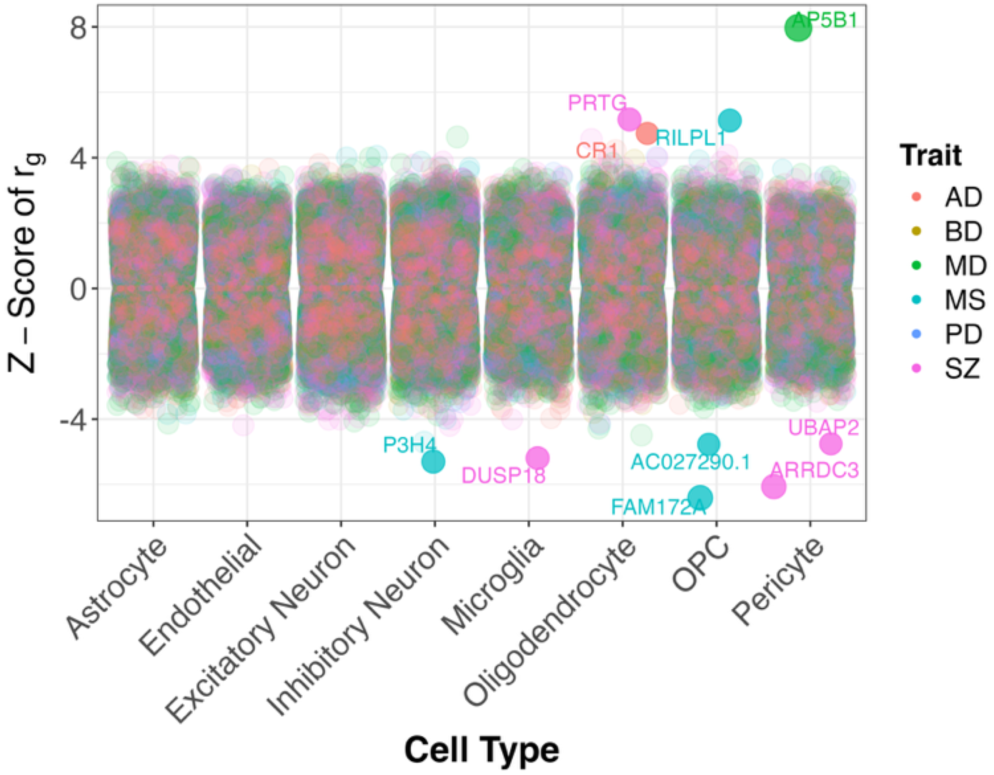
**Disorder-gene correlations using single cell eQTL data**. The local genetic correlation between SNP effect for a given gene expressed in a cell type (x-axis) is depicted by a dot. The strength of the local genetic correlation is expressed as a z-score (y-axis).

## Discussion

In this study we present and validate bivariate GSA-MiXeR, a novel functional genomics tool that partitions heritability and covariance for two traits within pre-defined genomic ROIs. A novel feature of our tool, that was supported by simulations, is the accurate estimation of the local r_g_ in disjoint genomic ROIs making our tool free from the restriction to continuous LD-based regions. In addition, through the LoGO statistic we provide a new metric to rank genomic ROIs based on its bivariate importance. In our applied analyses, we partitioned heritability across cell-type gene sets for six brain-related disorders and observed patterns of enrichment consistent with previous reports. We also partitioned covariance among those disorders across the cell-type gene sets to reveal local r_g_ associations not previously feasible. We showed that the LoGO statistic provides alternative bivariate information to the local r_g_ where the former highlights genomic ROIs and the latter pairs of disorders when performing cluster analyses. Through extended analyses we also illustrate the utility of bivariate GSA-MiXeR to inform individual-level genetic liability to disorders and to implicate cell-specific genes. Overall, bivariate GSA-MiXeR is a powerful tool for exploring local genetic associations within biologically relevant genomic ROIs for pairs of complex human traits.

Our cell type analyses illustrate the utility of bivariate GSA-MiXeR to replicate known cell type enrichments for brain-related disorders and highlight putative genomically regulated neural cell mechanisms. In line with previous reports, the psychiatric disorders were enriched for several neuronal cell types, however, the pattern of enrichment was distinct to a disorder.^16,17,58^ Meanwhile, each neurodegenerative disorder exhibited a unique pattern of cell type enrichment with AD and MS sharing enrichment for immune cell types consistent with previous reports.^16,59^ The PD enrichment for cell types sampled from the early prenatal brain may suggest that early brain development can play an important role for this neurodegenerative disorder or mechanisms disrupted by the disorder have key functional roles in early development. The former is supported by human midbrain like organoids studies which report that PD-related genetic mutations result in neurodevelopmental defects and transcriptomic alterations;^60,61^ while, the latter is supported by studies of fetal neural cell and induced pluripotent stem cell transplantation for the treatment of PD which improves symptomology.^62–64^ Overall, we replicate previous reports of cell type enrichment for these disorders and highlight important biological features that can have clinical utility.

By performing local r_g_ analyses within biologically relevant genomic ROIs, bivariate GSA-MiXeR can highlight putatively shared mechanisms. Here, we report the first local r_g_ estimates between pairs of psychiatric and neurological traits measured within neural cell-type gene sets. We observed pervasive local genetic overlap among the psychiatric disorders, putative astrocyte involvement for neurodegenerative disorders, and highlighted many cell types that may play a role in the comorbidity of psychiatric and neurodegenerative disorders with oligodendrocyte-related cell types featured prominently. Two main novel findings stand out: (i) some psychiatric and neurodegenerative disorders had negative correlations within specific cell types and (ii) some cell types were implicated as both enriched and correlated between pairs of disorders. The presence of a significant negative correlation between two traits in a given cell type gene set may suggest an opposing genomically-regulated mechanism underlying the two disorders. One example is the negative r_g_ observed between BD and PD within the Lamp5-Lhx6 cortical neuron gene set. Interestingly, a snRNA-seq study of orbitofrontal cortex samples from adults aged 26-84 reported Lamp5-Lhx6 interneurons exhibited the greatest number of differentially expressed genes with aging.^65^ Coupled with the elevated risk of developing PD among BD patients,^66^ it is possible that Lamp5-Lhx6 cortical neurons may have opposing expression profiles and/or pathophysiological effects in these two disorders as a function of aging. We highlighted a list of cell types both enriched for all three psychiatric disorders and with correlated effects among all pairs of those disorders providing a novel illustration of putative pleiotropic effects which may offer important insights into shared mechanisms. Across clinical domains, MD and MS were both enriched and correlated for the MGE interneuron gene set. Notably, somatostatin expressing interneurons which arise from the MGE have been implicated in the pathophysiology of MD meanwhile loss of inhibitory neurons from the MGE lineage is also observed in MS.^67–70^ This shared neuronal mechanism may be a putative biological link underlying comorbidity. Our findings illustrate that bivariate GSA-MiXeR provides a novel avenue to assess shared mechanisms and understand the underlying biology of clinically overlapping and comorbid disorders.

There are some limitations to consider. Similar to univariate GSA-MiXeR, our bivariate extension does not provide formal p-values for fold enrichment (see the Methods). Coupling fold enrichment estimates with other tools such as MAGMA, as done previously,^3,71^ can provide corroborating evidence and p-values.^72^ Notably, MAGMA and some other approaches to gene set analyses do not identify significant depletion of genetic signal. Currently, bivariate GSA-MiXeR only models autosomes and relies on an LD reference of European ancestry. Future developments will extend our tools utility to include the X chromosome and the inclusion of additional genetic ancestries. For our applied analyses, we classify MS as a neurodegenerative disorder. We note that neurodegeneration is a feature of MS, but the disorder is not considered a classic neurodegenerative disorder as is AD and PD. Also, we refer to various cell types from differing data sources and stages of life as distinctive entities. It is important to note that these classifications more accurately represent cell states and that differing sources of data may refer to the same or similar cell state using differing terminology.

The utility of bivariate GSA-MiXeR far exceeds local gene set analyses. We illustrated how the univariate and bivariate estimates may be translated to inform the partitioning of individual-level liability to disorders using PRS. Such PRS analyses can be used to understand comorbidity at the individual level and inform disorder subtyping with important implications for targeted treatment. We also show how local r_g_s can be conducted at finer granularity to implicate specific genes using sc-eQTL data. Future studies can apply our method to investigate other genomic ROIs and differing GWAS traits. Overall, bivariate GSA-MiXeR enhances the ability to identify shared and distinct biological mechanisms underlying complex human traits.

## Resource Availability

### Data availability

The GWAS summary statistics used AD, BD, MD, SZ are publicly available from the PGC (https://pgc.unc.edu/for-researchers/download-results/), for MS from the IMSGC (https://imsgc.net/?page_id=31) and for PD from the International Parkinson’s Disease Genomics Consortium (https://bit.ly/2ofzGrk). The additional PD GWAS meta-analysis conducted for the PRS analyses is available on the GWAS catalog (Link TBD). The single cell RNA-seq data were all derived from studies from the National Institute of Healths Brain Research through Advancing Innovative Neurotechnologies (BRAIN) Initiative – Cell Census Network (BICCN). Data for the early prenatal sample was downloaded from here https://cellxgene.cziscience.com/collections/4d8fed08-2d6d-4692-b5ea-464f1d072077 and the adult brain sample was downloaded from here https://cellxgene.cziscience.com/collections/283d65eb-dd53-496d-adb7-7570c7caa443. The TOP and DemGene cohort data have controlled data access. These data are not publicly available due to national data privacy regulations as they contain information that could compromise research participant privacy and/or consent. The LD reference can be downloaded from the original GSA-MiXeR tool repository here https://github.com/precimed/mixer.

## Code Availability

The bivariate GSA-MiXeR tool with example code is available on GitHub (link provided upon publication). All additional tools are publicly available including MiXeR-Pred (https://github.com/precimed/mixer-pred); HDL-L (https://github.com/zhenin/HDL); PLINK (v1: https://www.cog-genomics.org/plink/; v2: https://www.cog-genomics.org/plink/2.0/); simu tool (https://github.com/precimed/simu); and PRSet (https://choishingwan.github.io/PRSice/).

## Data Availability

The GWAS summary statistics used AD, BD, MD, SZ are publicly available from the PGC (https://pgc.unc.edu/for-researchers/download-results/), for MS from the IMSGC (https://imsgc.net/?page_id=31) and for PD from the International Parkinson's Disease Genomics Consortium (https://bit.ly/2ofzGrk). The additional PD GWAS meta-analysis conducted for the PRS analyses is available on the GWAS catalog (Link TBD). The single cell RNA-seq data were all derived from studies from the National Institute of Healths Brain Research through Advancing Innovative Neurotechnologies (BRAIN) Initiative - Cell Census Network (BICCN). Data for the early prenatal sample was downloaded from here https://cellxgene.cziscience.com/collections/4d8fed08-2d6d-4692-b5ea-464f1d072077 and the adult brain sample was downloaded from here https://cellxgene.cziscience.com/collections/283d65eb-dd53-496d-adb7-7570c7caa443.
The TOP and DemGene cohort data have controlled data access. These data are not publicly available due to national data privacy regulations as they contain information that could compromise research participant privacy and/or consent. The LD reference can be downloaded from the original GSA-MiXeR tool repository here https://github.com/precimed/mixer.

https://imsgc.net/?page_id=31

https://bit.ly/2ofzGrk

https://cellxgene.cziscience.com/collections/4d8fed08-2d6d-4692-b5ea-464f1d072077

https://cellxgene.cziscience.com/collections/283d65eb-dd53-496d-adb7-7570c7caa443

https://github.com/precimed/mixer

## Acknowledgments

We want to acknowledge the Norwegian registry of persons assessed for cognitive symptoms (NorCog), patients and clinicians for providing access to patient data and biological material. Funding was provided by the following resources: the Research Council of Norway [grants 223273, 248778, 324252, 324499, 326813, 334920, 344121], the South-East Regional Health Authority [grant 2022-073, 2022-087, 2025-039,], EEA and Norway [grant EEA-RO-NO-2018- 0573], European Union’s Horizon 2020 Research and Innovation Programme [grant 964874], and the National Institutes of Health [grants NIH 5R01MH124839-02, U24DA041123, R01AG076838, U24DA055330, OT2HL161847]. This work was performed on resources provided by UNINETT Sigma2 (the National Infrastructure for High Performance Computing and Data Storage in Norway) and services from sensitive data (TSD; Tjeneste for Sensitive Data) facilities, University of Oslo Norway.

## Author Contributions

Conceptualization: NP, OF, AS, AMD, OA; Data curation: NP, OF, AS, JF, DN, VF, SB, EH, DA, SB, TE, TF, KP, AR, IS, SBS, KS, GS, ES, MT, LW; Formal analysis: NP, OF, JF, DN AS; Funding acquisition: OAA, AMD; Software: NP, OF, JF, DN; Visualization: NP, OF, JF; Writing – original draft: NP, OF; Writing – review & editing: all listed authors.

## Methods

### General Additive Model

In an additive genetic model, a random trait 𝑦 ∈ ℝ is modeled as a linear combination of genotype 𝑔_𝑖_ with effect size 𝛽_𝑖_ as follows:

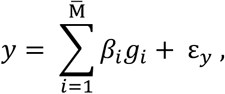

where 𝑖 iterates over Μ̄ variants, and ε_𝑦_ is the normally distributed 𝒩(0,1 − ℎ^2^) residual error including environmental effects. Effect sizes 𝛽_𝑖_ ∈ ℝ represent additive genetic effects and are random variables. Genotypes 𝑔_𝑖_ are centered but not scaled, 𝑔_𝑖_ ∈ {0 − 2𝑓_𝑖_, 1 − 2𝑓_𝑖_, 2 − 2𝑓_𝑖_}, where 𝑓_𝑖_ is the allele frequency of the 𝑖 − 𝑡ℎ SNP. The trait 𝑦 is centered and scaled such that 𝐸[𝑦] = 0, Var[y] = 1, thus the variance Var[∑_𝑖_ 𝛽_𝑖_𝑔_𝑖_] the traits SNP heritability, ℎ^2^ ∈ [0,1].

With bivariate GSA-MiXeR we extend the general additive model to include priors on effect sizes, account for heterozygosity, include functional annotations, and we introduce a bivariate extension that partitions covariance in genomic ROIs.

### Model Estimation from GWAS Z-Scores

Consistent with the original GSA-MiXeR,^3^ we infer model parameters using GWAS z-scores (𝑧_j_) with the index 𝑗 = 1, … , 𝑀 iterating over GWAS tag SNPs that intersect with the genomic LD reference panel (𝑀). GWAS-based 𝑧_j_ is related to the additive genetic effects of the general model 𝛽_𝑖_ as follows:

Let 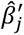 be GWAS SNP estimates of marginal effects for the 𝑗 − 𝑡ℎ tag SNP and 𝑧_j_ be the corresponding z-score where 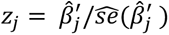 Then,

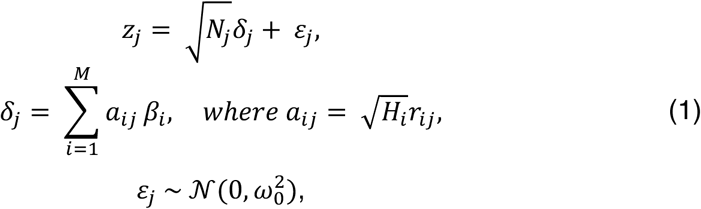

where 𝑁_j_ is the number of GWAS participants with non-missing genotype for the 𝑗 − 𝑡ℎ SNP. 𝐻_𝑖_ = 2𝑓_𝑖_(1 − 𝑓_𝑖_) denotes the heterozygosity of the 𝑖-th SNP. 𝑟_𝑖j_ is the LD correlation coefficient between genotype vectors of the i-th and j-th SNP. The parameter ^2^ accounts for non-polygenic inflation in GWAS z-scores where 𝜀_j_denotes a normally distributed residual. The proof of the above equations can be found in the Supplementary Methods

### Fold Enrichment of Heritability

Within our bivariate GSA-MiXeR framework, we introduce three main models for the estimation of univariate and bivariate outputs: (i) a null model, which incorporates MAF-dependent genetic architecture; (ii) a baseline model, which builds on the null model to account for non-uniform heritability distribution across functional annotations; and (iii) a full model, which builds on the baseline model to account for genomic ROIs. Details on the complete notation of these models can be found in the Supplementary Methods. To estimate fold enrichment 𝑓(𝐺) for a specific genomic ROI, 𝐺, we fit parameters of the full model while keeping the baseline model parameters fixed, and define 𝑓(𝐺) such that:

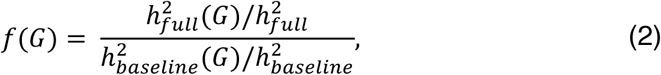

where 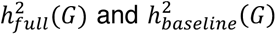 denote the heritability estimates from the full or base model using SNPs within the genomic ROI, respectively. Meanwhile, 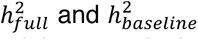 denote the genome-wide heritability estimates from the full or the base model, respectively. The GSA-MiXeR log likelihood function for observing 𝑧_j_given the modeled parameters can be found in the Supplementary Methods. We note that fold enrichments can be estimated using full vs null models as well (i.e., not accounting for functional annotations) which may be useful for several implementations of the tool including if those functional annotations are to be modelled as genomic ROIs.

### Local Genetic Correlation and Local Genetic Omnibus Statistic

In our estimation of bivariate parameters, we model the heritability of each trait separately based on the null, baseline, or full model. The genetic correlation between two traits is defined as the covariance of their genetic components, normalized by the square root of the product of the heritability of both traits: 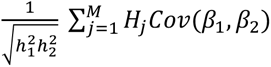, where 𝐻_j_ is the heterozygosity of the 𝑗-th variant. The same equation constrained to SNPs in a given genomic ROI defines local r_g_. The full bivariate model allows correlation parameters (𝜌_𝐺_) in 𝐶𝑜𝑣(𝛽_1j_, 𝛽_2j_’ = 𝜌_𝐺_𝜎_1j_𝜎_2j_to be estimated separately for each genomic ROI. A maximum likelihood optimization procedure is based on the following bivariate distribution for z-scores:

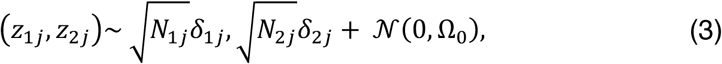

extending univariate model (1) and incorporating the cross-trait intercept 𝜌_0_𝜔_01_𝜔_02_ in the residual term 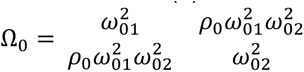. Details can be found in the Supplementary Methods along with the full expression of the log-likelihood function for estimating these parameters. We generate likelihood-based p-values for the local r_g_ estimates using a likelihood ratio test that compares the full model and a reduced model which holds the local r_g_=0.

The LoGO statistic is defined as the difference in log-likelihood between the bivariate full and baseline model. Therefore, the heritability of both traits and their local r_g_ within the genomic ROI is modeled in the bivariate full model while the baseline model is agnostic to the genomic ROI.

### LD Reference Panel

The Haplotype Reference Consortium (HRC) is used as our default LD reference panel in line with the original GSA-MiXeR.^3^ Sample quality control (QC) for generating the HRC reference included selection of individuals with European ancestry and removal of related individuals (i.e., up to second degree relatives) resulting in 23,152 samples included. For SNP QC, we excluded SNPs with missing or duplicated RS IDs and used the following PLINK settings -- maf 0.001 --hwe 1e-10 --geno 0.1 which resulted in the inclusion of 11,980,511 SNPs in the reference. For the HRC reference, the LD r2 was calculated for each chromosome separately using a window of 10MB and a minimum r2 of 0.01. All analyses with bivariate GSA-MiXeR described in this paper used this HRC reference restricted to HapMap3 SNPs (n=1,117,340).

### Simulations

We used UK Biobank data for all simulations. Here sample QC involved selection of individuals of white British ancestry, exclusion of participants with a sex chromosome aneuploidy, and removal of participants with withdrawn consent to use data which resulted in 337,145 individuals. SNP QC involved the exclusion of SNPs with duplicated RS IDs, excluded SNPs with an imputation INFO score below 0.8, and used the following PLINK arguments –maf 0.001 --hwe 1e-10 --geno 0.1 which resulted in a total of 12,926,669 SNPs included. After QC, we selected a random subset of 100,000 unrelated individuals. In addition, all analyses conducted using bivariate GSA-MiXeR by default excludes variants in the MHC region.

To simulate pairs of quantitative traits we used the simu tool (https://github.com/precimed/simu) which samples causal effects from a bivariate normal distribution. For each pair of traits, we simulated equal polygenicity (𝜋 = 0.003, 𝑛 = 3296 SNPs) and heritability (ℎ^2^ = 0.3). Each pair of traits was also simulated to share 50% of their causal SNPs where the genetic correlation in this shared component varied taking on one of the following nine values: −1, −0.75, −0.5, −0.25, 0, 0.25, 0.5, 0.75, 1. Therefore, simulations included three sets of causal SNPs, two trait-specific and one shared set. For each simulated trait, SNP effects were sampled from a bivariate normal distribution and scaled to reach a set heritability (ℎ^2^ = 0.3). Then for each of the 100,000 UK Biobank participants, a trait was generated as the sum of allelic effects along with environmental effects modelled as random Gaussian noise. GWAS were then performed using PLINK2^73^ adjusting for age, sex, and the first 10 genetic principal components.

Similar to bivariate GSA-MiXeR, the HDL-L tool uses a random effects model and likelihood-based estimation of local genetic correlations. Therefore, we performed additional simulations to compare the two methods. The local genomic ROIs were defined using the 34 continuous LD-based genomic regions on chromosome 22 generated by the creators of HDL-L.^6^ We simulated pairs of quantitative traits with equal polygenicity (𝜋 = 0.003, 𝑛 = 3296 SNPs) and heritability (ℎ^2^ = 0.3) and with 50% of their causal SNPs shared as described above. For each genomic ROI, 5 pairs of traits were simulated at varying genome-wide r_g_ −1 and 1 by 0.25 increments. This resulted in 45 simulated pairs of traits for each LD-based genomic ROI. For each pair of traits, 20% of the SNPs contained in the genomic ROI were randomly selected to be causal. The rest of the causal SNPs were randomly selected from SNPs that were not contained within the genomic ROI. Both GSA-MiXeR and HDL-L were run for all 45 trait pairs for each of the 34 continuous genomic ROIs (n=1,530 simulated trait pairs). The estimated local r_g_ from both methods was then compared to the true correlation in causal betas using a Pearson correlation and mean squared error estimates (MSE) were compared between methods.

To assess bivariate GSA-MiXeR performance when applied to genomic ROIs defined by gene sets, we used a set of 18,201 protein-coding genes consistent with our original GSA-MiXeR tool. These genes were randomly assigned to belong to one of three groups: trait 1, trait 2, or shared. Then, for each gene, SNP assignment was performed using a 10kbs window around the gene. The resulting set of three SNP lists (i.e., trait 1, trait 2, or shared) restricted to protein-coding genes was used as a pool of putative causal SNPs to draw from in simulations. During simulations, a random set of causals was drawn from these lists of SNPs to model the two-trait specific and one shared component. For each of the nine simulated genetic correlation values (−1, −0.75, −0.5, −0.25, 0, 0.25, 0.5, 0.75, 1), we generate 10 random pairs of traits using these lists of causal SNPs. After the pairs of traits were simulated, the shared causal SNPs for a given pair of traits were used to generate gene sets specific to the shared component of those traits by mapping the randomly selected causals SNPs back to their respective genes. Then, we generate gene sets of varying size (10, 25, 50, 100, 250, 500, or 1000 genes). These gene sets are randomly generated from the list of shared genes with 20 iterations per gene set size. We then run bivariate GSA-MiXeR for each of the 90 trait pairs and their related 140 gene sets. We compare the true local r_g_ (calculated across causal SNPs) with the estimated local r_g_ by bivariate GSA-MiXeR using a Pearson correlation.

The final null simulations were constructed to validate the p-values for the local r_g_ estimates. Here, each pair of traits was simulated to have 100% overlap and therefore one causal component made up a single list of shared SNPs. The same polygenicity (𝜋 = 0.003, 𝑛 = 3296 SNPs) was modeled as above and the genome-wide r_g_ was set to zero between the simulated trait pairs. All causal SNPs were drawn from a randomly generated gene set made up of 100 protein-coding genes across 1000 repetitions. Since trait heritability was now restricted to this causal gene set, we varied these values as follows: ℎ^2^ = 0.0001, 0.0005 𝑜𝑟 0.001. We then ran bivariate GSA-MiXeR for each of the 3,000 traits and their specific causal gene set.

### GWAS Summary Statistics

We used summary statistics from the latest psychiatric genomics consortium GWAS of BD^10^, MD^14^, and SZ^18^. We acquired summary statistics for AD from the psychiatric genomics consortium GWAS by Wightman et al (2021)^15^, for PD from the GWAS by Nalls et al (2019)^9^, and for MS from the international multiple sclerosis genetics consortium^20^. All GWAS data included participants of European ancestry only in alignment with the reference panel for bivariate GSA-MiXeR. To avoid sample overlap in the polygenic score analyses (see below), we used summary statistics excluding participants in the target sample for AD, BD, and SZ. In addition, to avoid overlap with our PD target sample for PRS analyses, we conducted a GWAS meta-analysis of non-overlapping samples. Specifically, we meta-analyzed the Nalls et al (2014) GWAS^74^ as well as the biobank samples contained in the recent meta-analysis from the Global Parkinson’s Genetics Program^75^. Applying the METAL tool (version “2020-05-05”) we performed a fixed-effects inverse-variance meta-analysis.

### Single Cell and Nucleus RNA-seq Data and Analyses

To derive cell-type gene sets we used three published datasets from the BRAIN initiative cell census network. We acquired gene expression data from (i) a scRNA-seq study of over 1.5 million cells extracted from early prenatal brain tissue of 26 donors,^76^ (ii) a snRNA-seq study of over 3 million nuclei extracted from cells sampled across the brains of 3 adult donors,^24^ and (iii) a snRNA-seq study of over 1.1 million nuclei extracted from cells sampled from the cortex of 5 adult donors.^23^ From the study of early prenatal brain, we defined cell types based on the annotation of 11 classes of cells. From the study of adult brain, cell types were defined using the annotations of 31 superclusters of cells. In line with previous reports, we use the top decile of expression proportion to generate cell type gene sets for the early prenatal and adult brain datasets.^16,17^ For a given cell type, the process involves calculating the average expression of each gene across cells, then for a given gene, calculating the expression proportion across cell types, and then selecting the top 10% of genes ranked with the highest proportion of expression for a given cell type. The resulting list of top expressed genes typifying a given cell type were used for all subsequent analyses. From the study of adult cortex, we used the published marker genes for the annotated 24 subclasses of cells and selected those consistently identified across the 8 cortical brain regions. The resulting cell type gene sets therefore consisted of marker genes conserved across cortical regions.

In our applied analyses we filter all cell type genes to be among a reference set of 18,201 protein coding genes from the original GSA-MiXeR tool^3^ and SNPs from the LD reference were assigned to a given gene using a 35kb upstream and 10kb downstream window around the gene. An average of 830.08 (sd=248.62, range=500-1347) genes for cell types from the early prenatal brain source, 1064.90 (sd=224.57, range=550-1548) genes for cell types from the adult brain source, and 87.25 (sd=97.29, range=8-320) genes for cell types from the adult cortex source were included in the gene sets for analyses. We then run bivariate GSA-MiXeR using all possible pairs of the six brain-related disorders and 68 cell type gene sets. In the case of AD, and in line with previous work, we remove chromosome 19 to avoid biases in modelling genetic architecture caused by the strong APOE effect.^8^ In line with the original GSA-MiXeR, our fold enrichment values have an associated standard error and delta AIC value but no p-values.^3^ Throughout the manuscript, we refer to a cell type gene set as enriched when the fold enrichment value is above one, the delta AIC value (based on the null, baseline, or full model comparisons) is above zero, and the fold enrichment plus or minus the standard errors does not cross one. Similarly, we refer to a cell type gene set as depleted when the fold enrichment is below one and the delta AIC value is above zero, and the fold enrichment plus or minus the standard errors does not cross one. This criteria is consistent with the exploratory analyses in the original GSA-MiXeR manuscript.^3^ We perform hierarchical clustering of fold enrichment values as well as the bivariate local r_g_ and LoGO statistics with the ComplexHeatmap package in R (v4.4.1) using the default Euclidian distance metric. Prior to clustering, fold enrichment values and LoGO statistics are scaled across cell types within a given disorder such that they are mean centered and have a standard deviation equal to one. We also use the z-score of the local r_g_ for clustering by taking the inverse of the cumulative distribution function of the p-values divided by two and assigning the sign of the local r_g_. LD score regression^77^ was used to estimate the genome-wide genetic correlation between all pairs of traits. The Benjamini-Hochberg method was used to correct p-values for multiple testing.

### Polygenic Risk Score Data and Analyses

Polygenic Risk Score (PRS) analyses were conducted using MiXeR-Pred which models the genetic architecture of an input trait (univariate) or pair of traits (bivariate) using z-scores from the input GWAS summary statistics.^52^ The first and second moments of the posterior distribution for both the univariate and bivariate models are used as new SNP weights and thresholding values in PRS estimation. To generate cell type PRS, we used the MiXeR-Pred weights and thresholding values as inputs for PRSet, a tool commonly used for pathway PRS analyses.^53^ PRSet input bed files (which contain coordinates) were generated for each gene set with a 35kbs upstream and 10kbs downstream window around each cell type gene to account for putative regulatory SNP effects.^53^

To generate PRS we used the DemGene sample for the neurodegenerative disorders AD and PD as well as the Thematically Organized Psychosis (TOP) sample for the psychiatric disorders BD and SZ. From the DemGene sample we included 2863 AD cases, 620 PD cases, and 1791 controls with an average age of 72.84 (sd=10.81) and 61.26% of participants were female. From the TOP sample we included 1070 BD cases, 1042 SZ cases, and 1422 controls with an average age of 35.57 (sd=13.81) and 47.22% of participants were female. To generate PRS in these samples, we used GWAS summary statistics that removed these cohorts resulting in no sample overlap. We excluded related individuals and QC of genetic data used the following PLINK arguments --maf 0.01 --hwe 1e-10 --geno 0.05.

The ability of bivariate GSA-MiXeR to partition heritability and perform local genetic associations within gene sets or pathways may inform pathway-specific polygenic risk scoring. Therefore, we conducted a series of association analyses at the univariate and bivariate level. We first used univariate MiXeR-Pred weights and thresholds to generate cell type PRS for AD, PD, BD, and SZ using PRSet. Separate linear models controlling for age, sex, genotyping batch and the first 20 genetic principal components were then used to test the association between a given cell type PRS and the associated disorder. For those cell type PRS – disorder associations that were significant after correction for multiple comparisons using the Benjamini-Hochberg method, we correlated the variance explained by those PRS with the fold enrichment estimates from bivariate GSA-MiXeR. At the bivariate level, we selected a secondary trait for each of AD, PD, BD, and SZ by using (i) the highest average absolute value in z-score of the local r_g_ across all cell types or (ii) the highest average LoGO statistic across all cell types. We then generated weights and thresholds from the bivariate MiXeR-Pred models and used those to create cell type PRS using PRSet. As with the univariate analyses, linear models controlling for age, sex, genotyping batch and the first 20 genetic principal components were then used to test the association between a given bivariate cell type PRS and the disorder associated with the primary trait. For those cell type PRS associations that survive correction for multiple comparisons (Benjamini-Hochberg method), we correlated the variance explained by those PRS with the (i) local r_g_ or (ii) LoGO statistic.

### Single Nucleus eQTL Data and Analyses

We used data from a study by Bryois et al (2022) which provided snRNA-seq eQTL data to implicate cell specific genes for each brain-related disorder.^78^ This study included 192 donors and derived eight brain cell types from samples of frontal/temporal cortex and white matter. The list of SNPs for a given gene within a given cell type was used to define the genomic ROIs. For each disorder, we iterated over all cell type genes and estimated the local r_g_ and associated p-values using bivariate GSA-MiXeR. For AD we again remove chromosome 19 to avoid biases in modelling genetic architecture caused by the strong APOE effect.^8^ The result is in a local r_g_ estimate between disorder genetic effects and cis-region genetic effects on gene expression within a given cell type. As with the sc- and snRNA-seq data above, we calculated z-scores for each disorder – gene pairing. The Benjamini-Hochberg method was used to correct p-values for multiple testing.

### Ethics Statement

Ethical approvals were obtained for all individual-level data used in our analyses. This data was conducted using UK Biobank data through application number 27412. For the UK Biobank, HRC, TOP, and DemGene samples approvals were obtained from the Norwegian Regional Committees for Medical and Health Research Ethics (REC; ref. 2009/2485, ref. 2014/631), Data Inspectorate (ref. 03/02051) and The Norwegian Directorate of Health (ref. 05/5821). The use of GWAS summary statistics was evaluated by the REC South-East Norway and deemed to not require additional ethical approval due to a lack of individual-level data (REC #2011/1980).

