## Supplementary Figures for "Bivariate GSA-MiXeR: A Novel Tool for Functional Genomic Analyses Implicates Diverse Neural Cell Types for Psychiatric and Neurodegenerative Disorders"

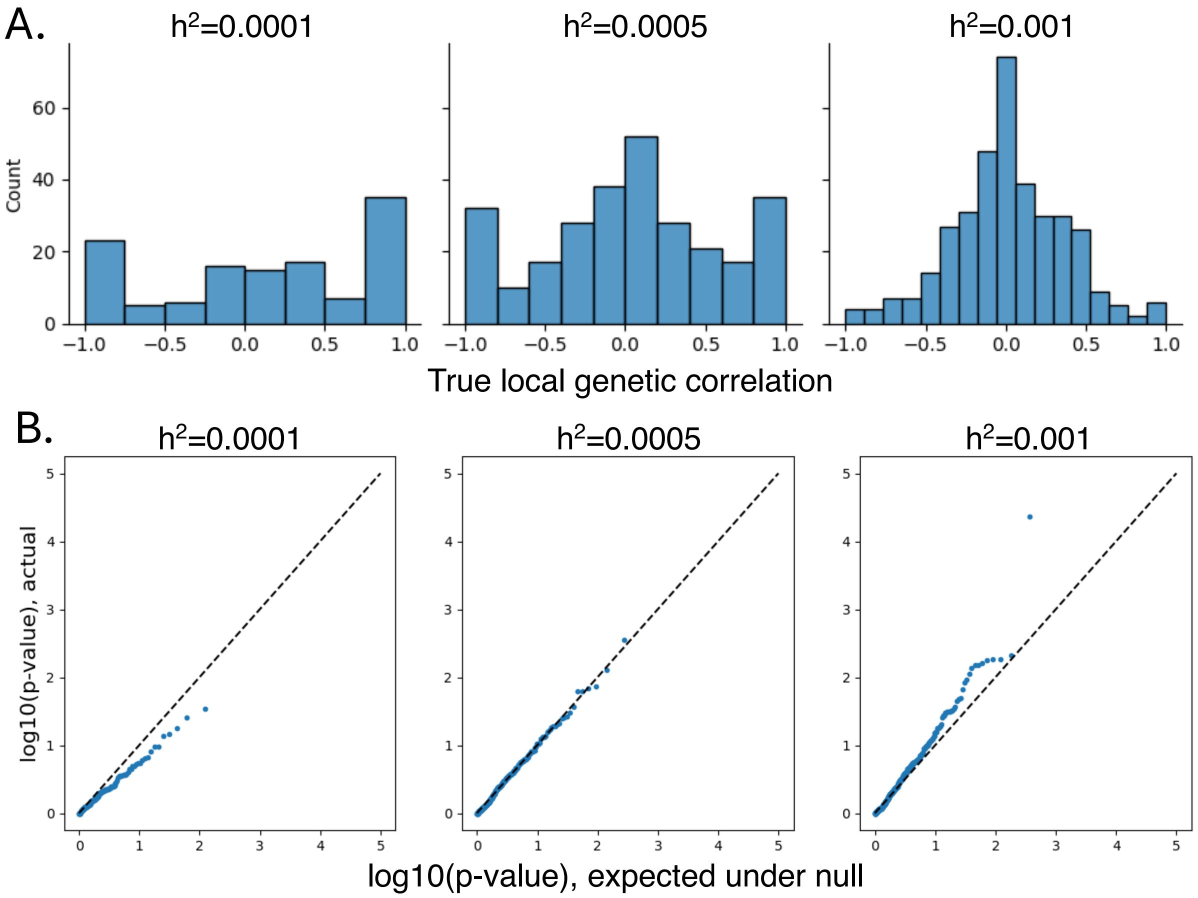


**Supplementary Figure 1. Null Simulations**. A) The distributions of local genetic correlations for null simulations using a random effects model and with fixed heritability values. B) Q-Q plots of the true and expected $\log_{10}$ p-values in the null simulations.

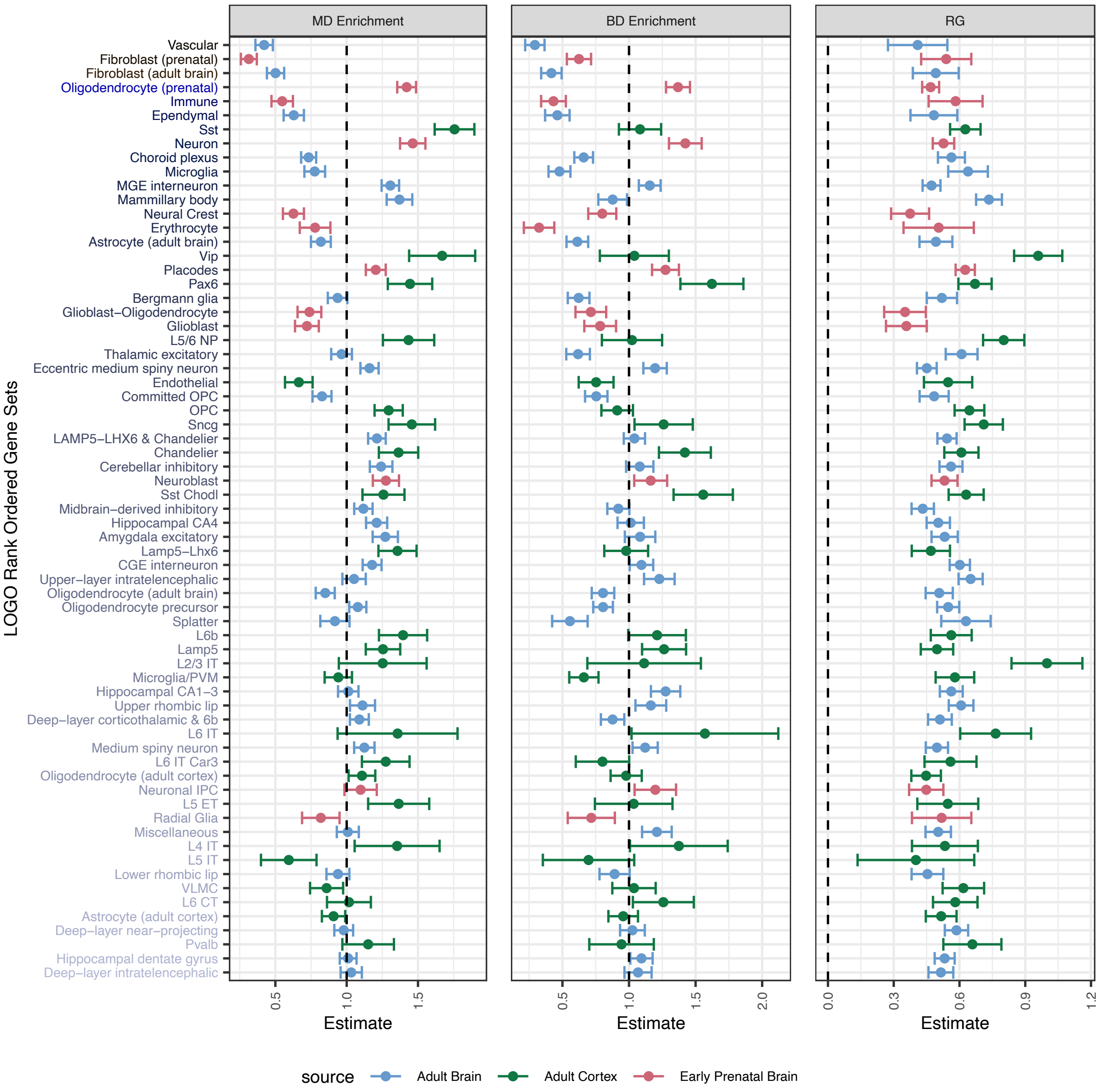


**Supplementary Figure 2. MD vs BD LoGO Rankings**. Depicted is the cell type gene sets on the y-axis ranked and colored based on the strength of the LoGO statistic. Each panel displays either the trait specific cell type enrichment or the local $r_g$ for a given cell type.

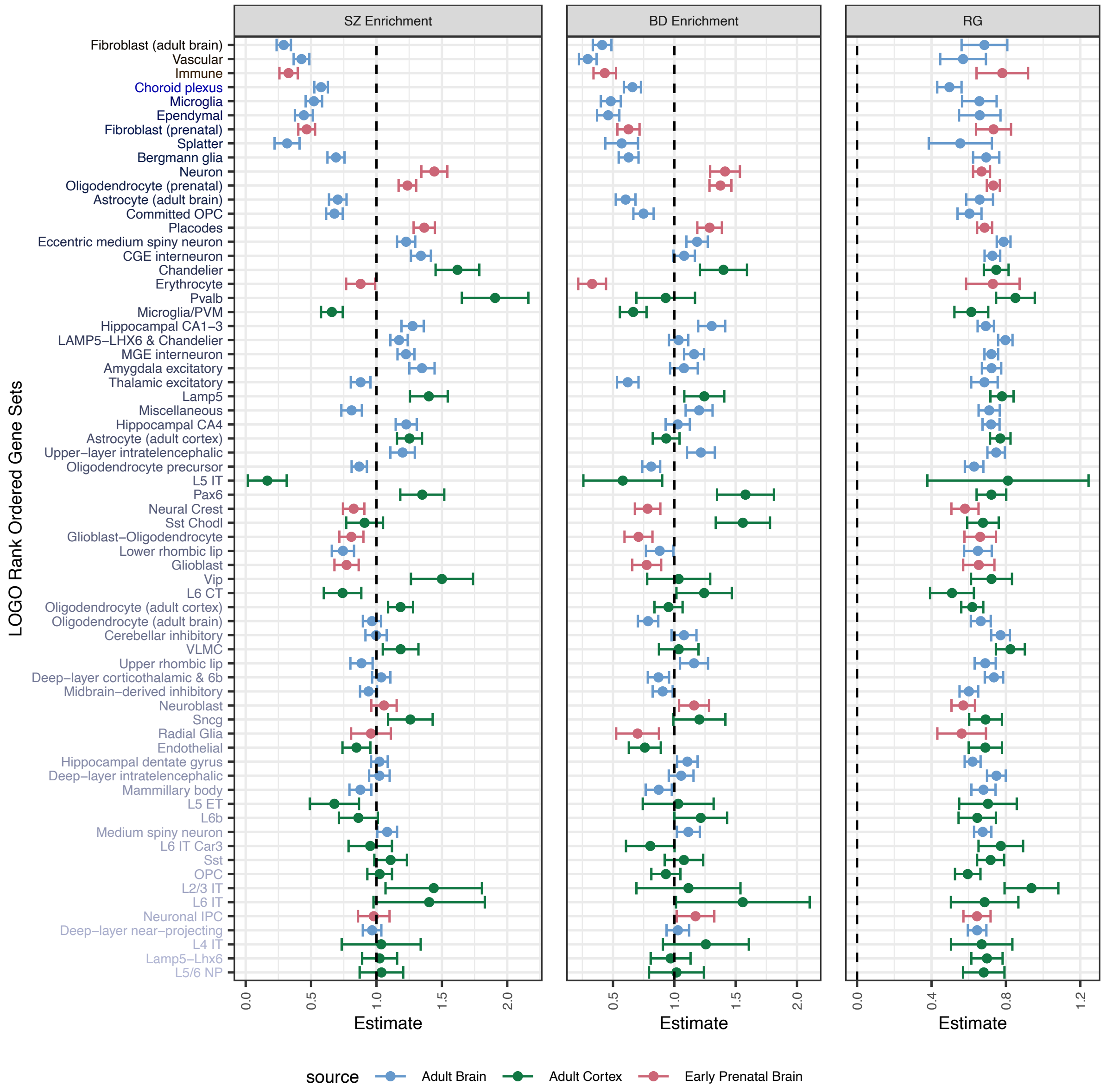


**Supplementary Figure 3. SZ vs BD LoGO Rankings**. Depicted is the cell type gene sets on the y-axis ranked and colored based on the strength of the LoGO statistic. Each panel displays either the trait specific cell type enrichment or the local $r_g$ for a given cell type.

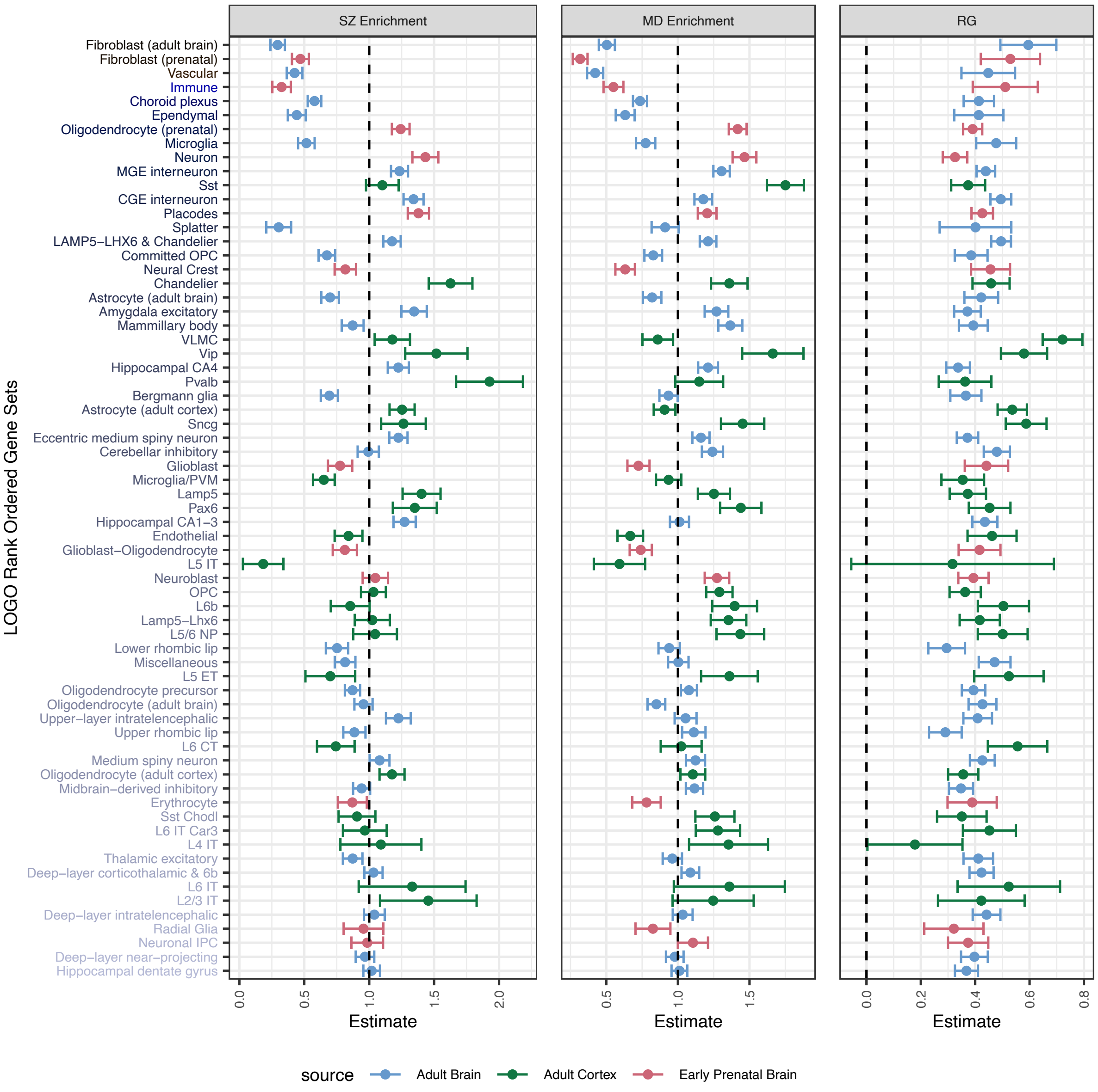


**Supplementary Figure 4. SZ vs MD LoGO Rankings**. Depicted is the cell type gene sets on the y-axis ranked and colored based on the strength of the LoGO statistic. Each panel displays either the trait specific cell type enrichment or the local $r_g$ for a given cell type.

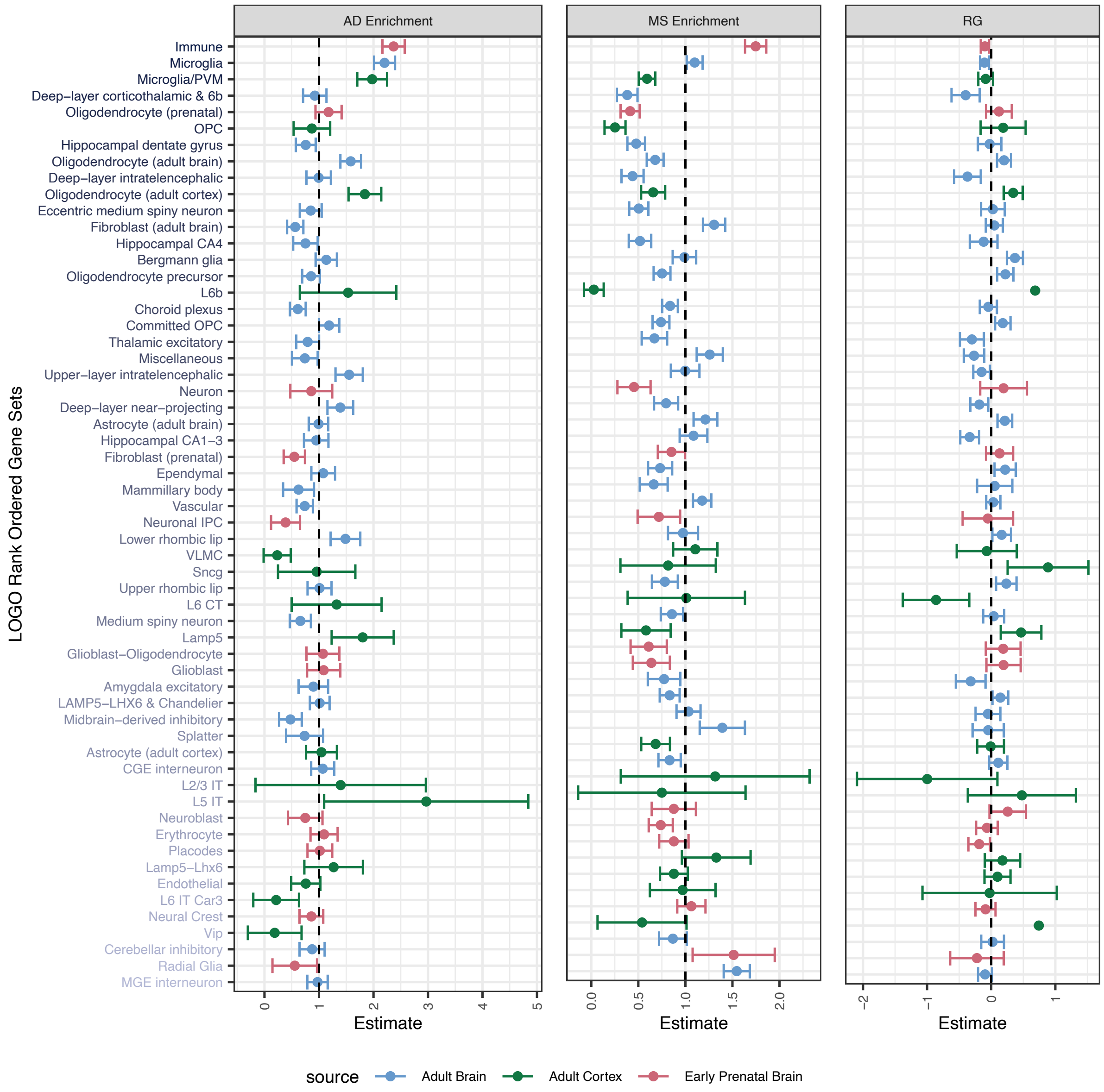


**Supplementary Figure 5. AD vs MS LoGO Rankings**. Depicted is the cell type gene sets on the y-axis ranked and colored based on the strength of the LoGO statistic. Each panel displays either the trait specific cell type enrichment or the local $r_g$ for a given cell type.

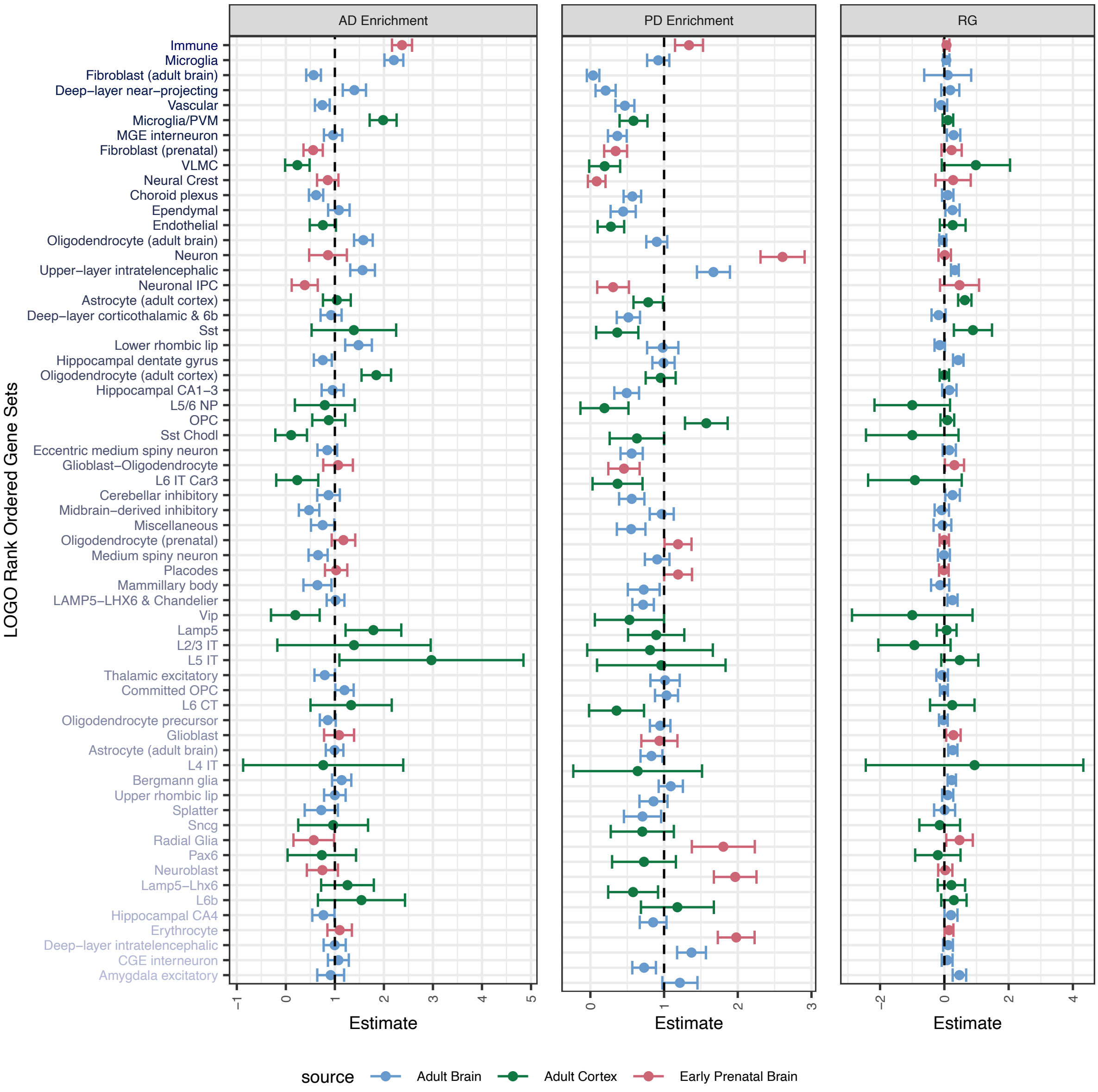


**Supplementary Figure 6. AD vs PD LoGO Rankings**. Depicted is the cell type gene sets on the y-axis ranked and colored based on the strength of the LoGO statistic. Each panel displays either the trait specific cell type enrichment or the local $r_g$ for a given cell type.

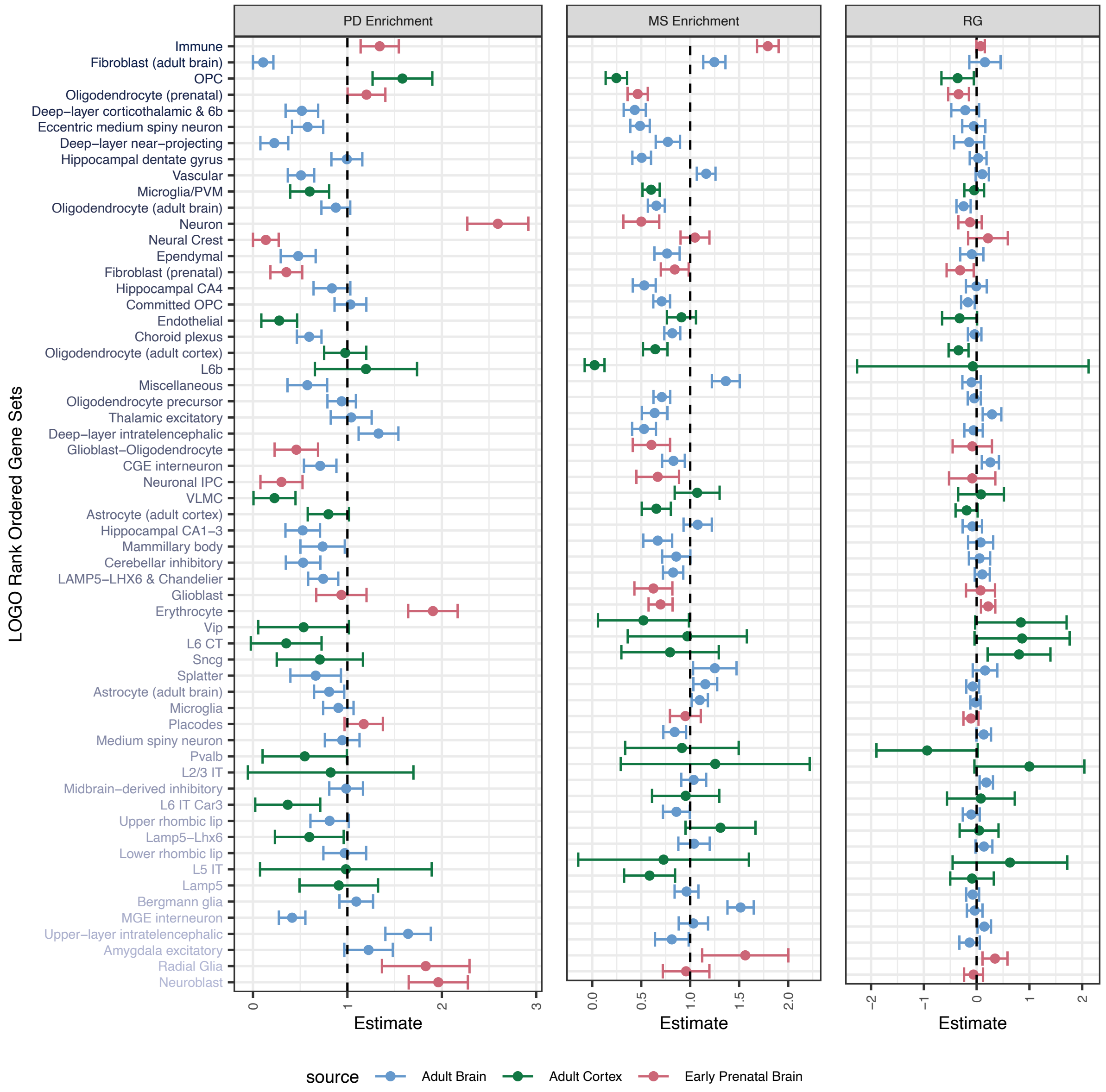


**Supplementary Figure 7. PD vs MS LoGO Rankings**. Depicted is the cell type gene sets on the y-axis ranked and colored based on the strength of the LoGO statistic. Each panel displays either the trait specific cell type enrichment or the local $r_g$ for a given cell type.

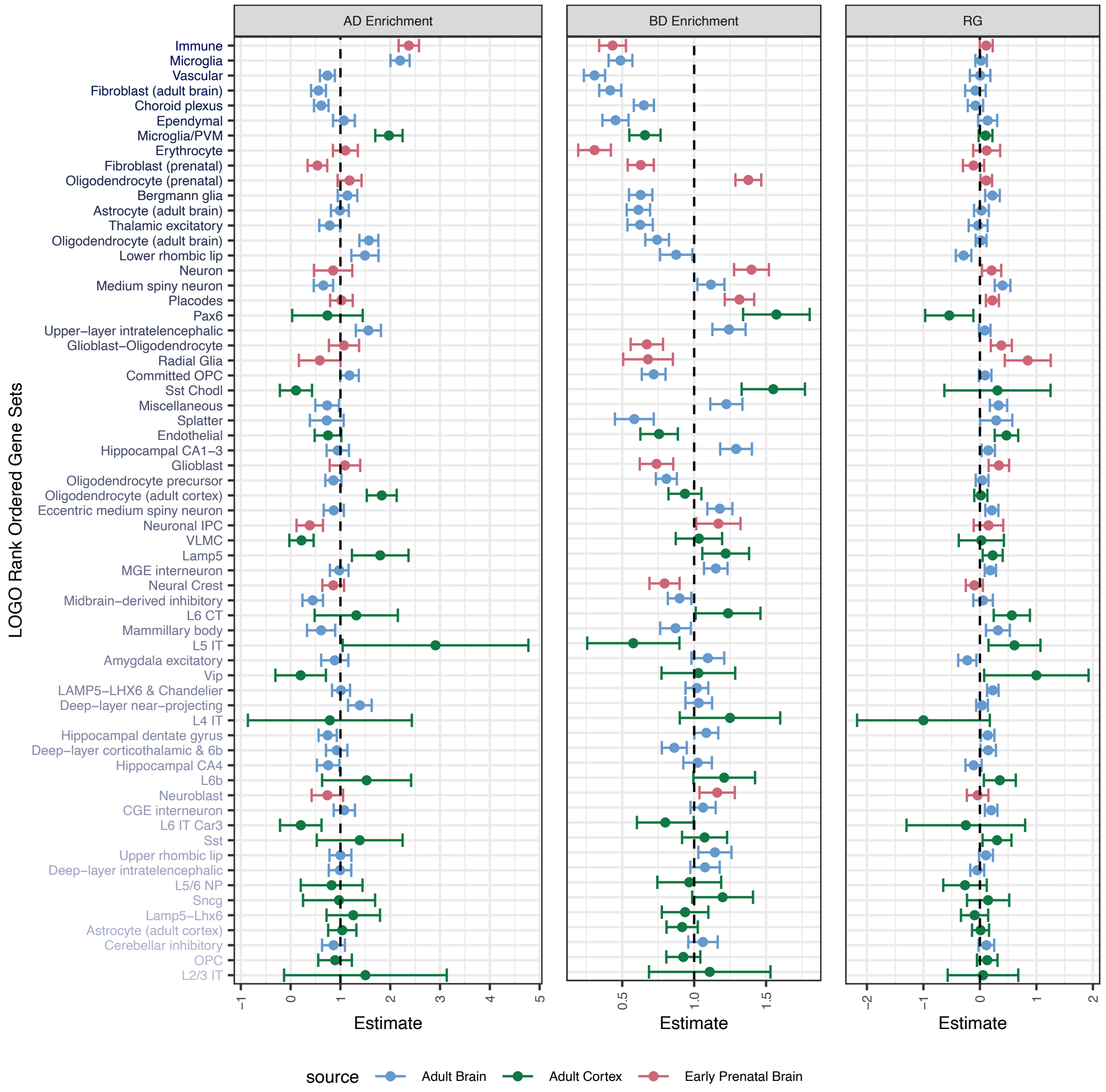


**Supplementary Figure 8. AD vs BD LoGO Rankings**. Depicted is the cell type gene sets on the y-axis ranked and colored based on the strength of the LoGO statistic. Each panel displays either the trait specific cell type enrichment or the local $r_g$ for a given cell type.

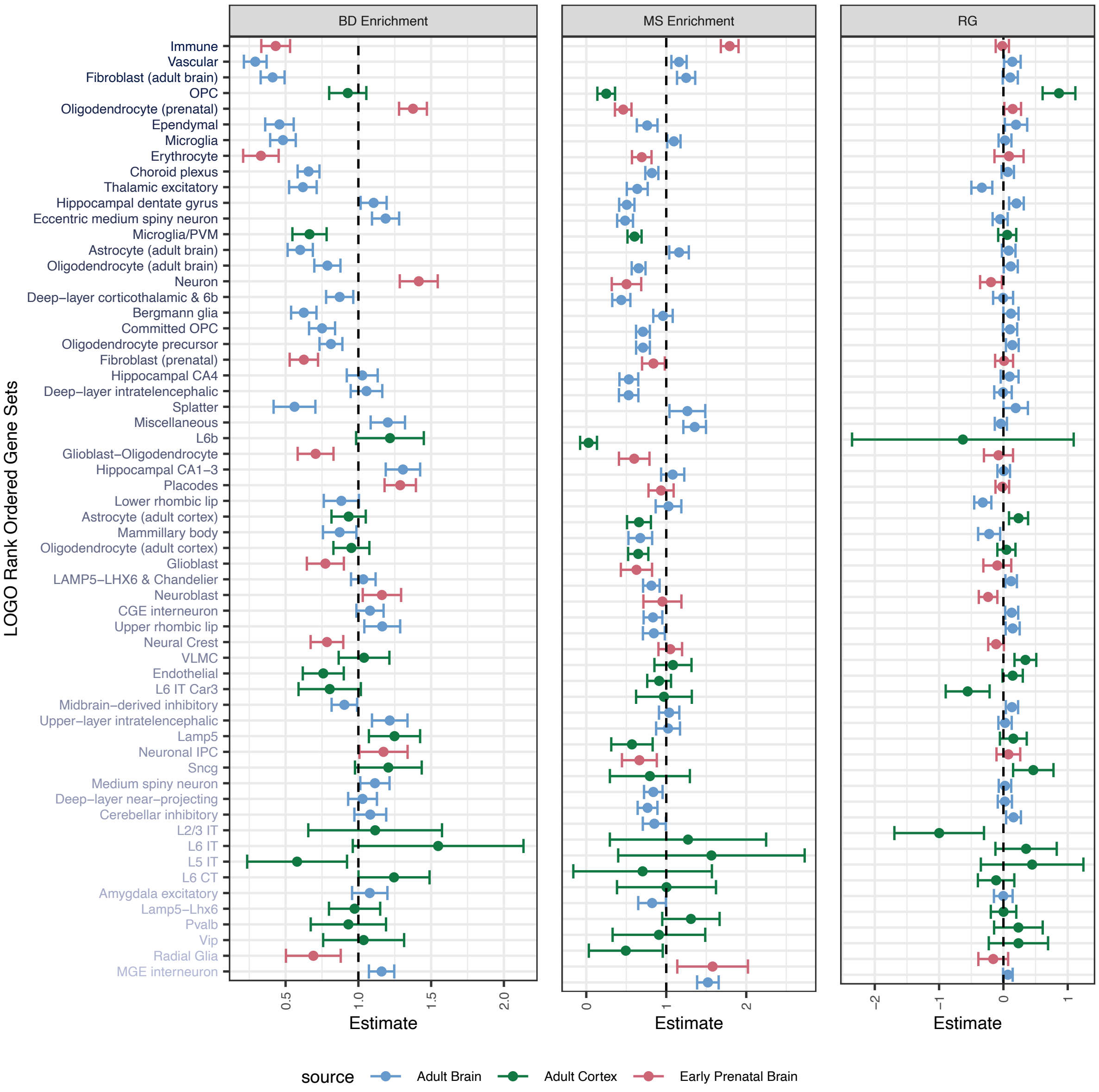


**Supplementary Figure 9. BD vs MS LoGO Rankings**. Depicted is the cell type gene sets on the y-axis ranked and colored based on the strength of the LoGO statistic. Each panel displays either the trait specific cell type enrichment or the local $r_g$ for a given cell type.

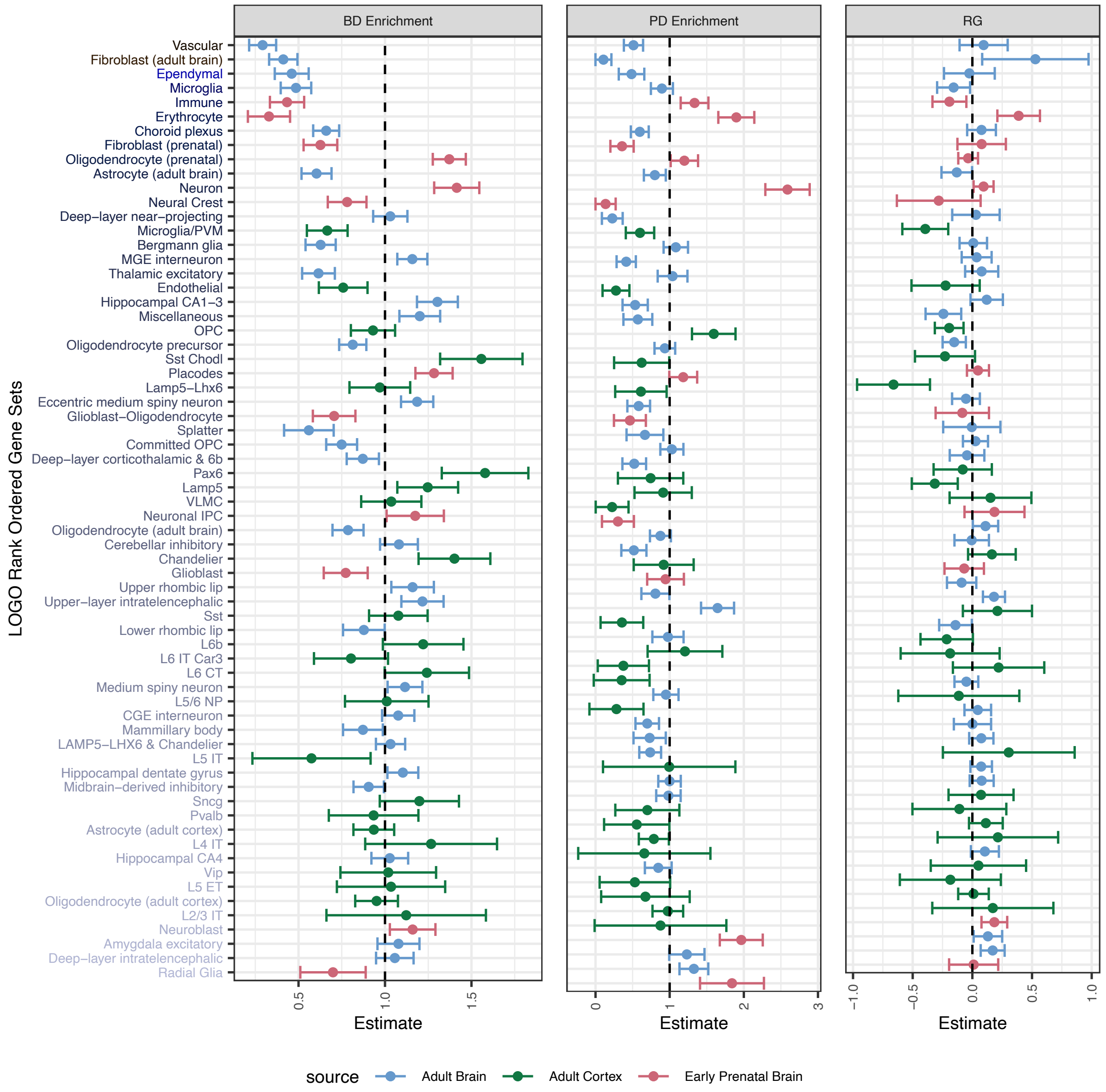


**Supplementary Figure 10. BD vs PD LoGO Rankings**. Depicted is the cell type gene sets on the y-axis ranked and colored based on the strength of the LoGO statistic. Each panel displays either the trait specific cell type enrichment or the local $r_g$ for a given cell type.

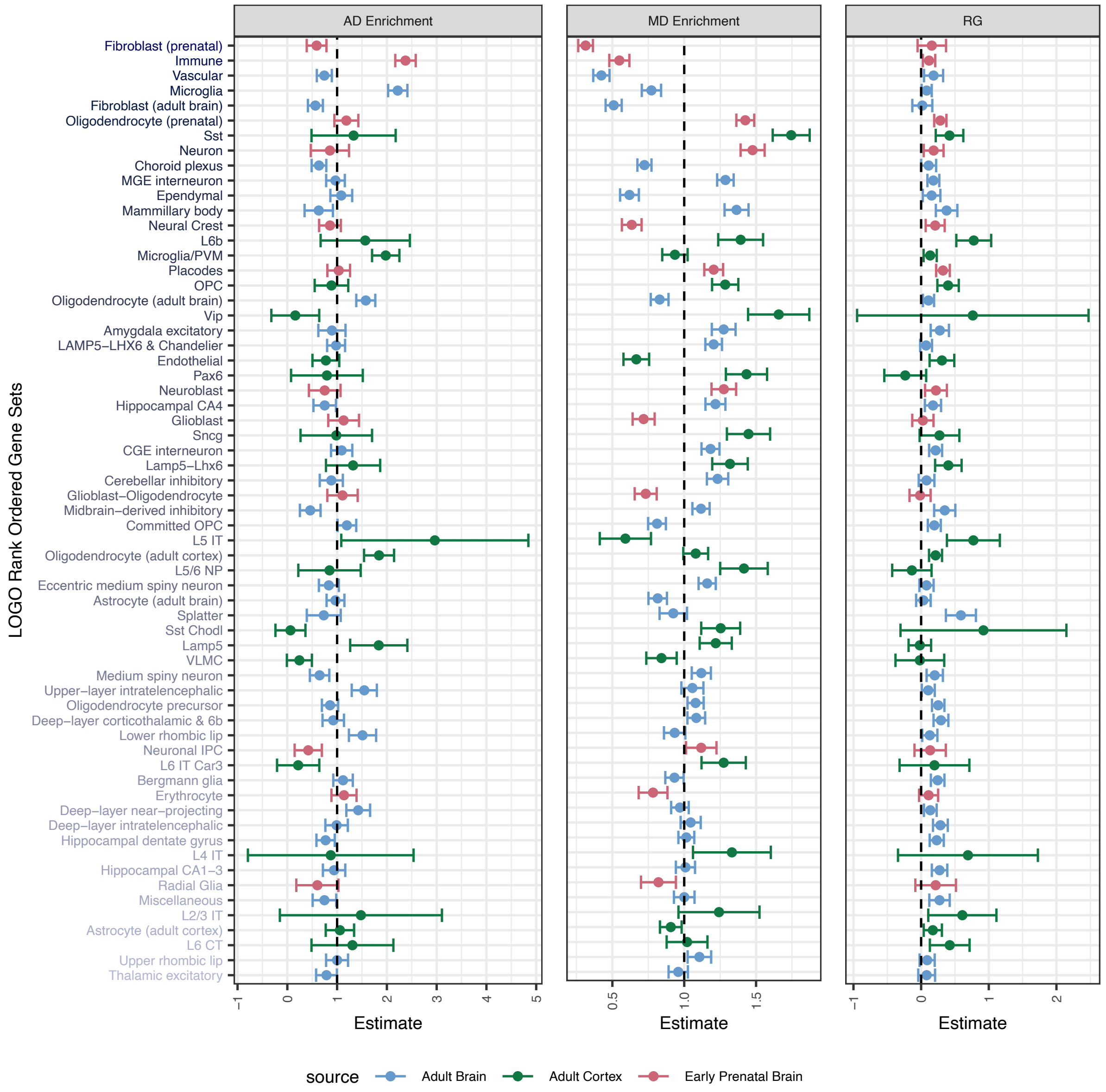


**Supplementary Figure 11. AD vs MD LoGO Rankings**. Depicted is the cell type gene sets on the y-axis ranked and colored based on the strength of the LoGO statistic. Each panel displays either the trait specific cell type enrichment or the local $r_g$ for a given cell type.

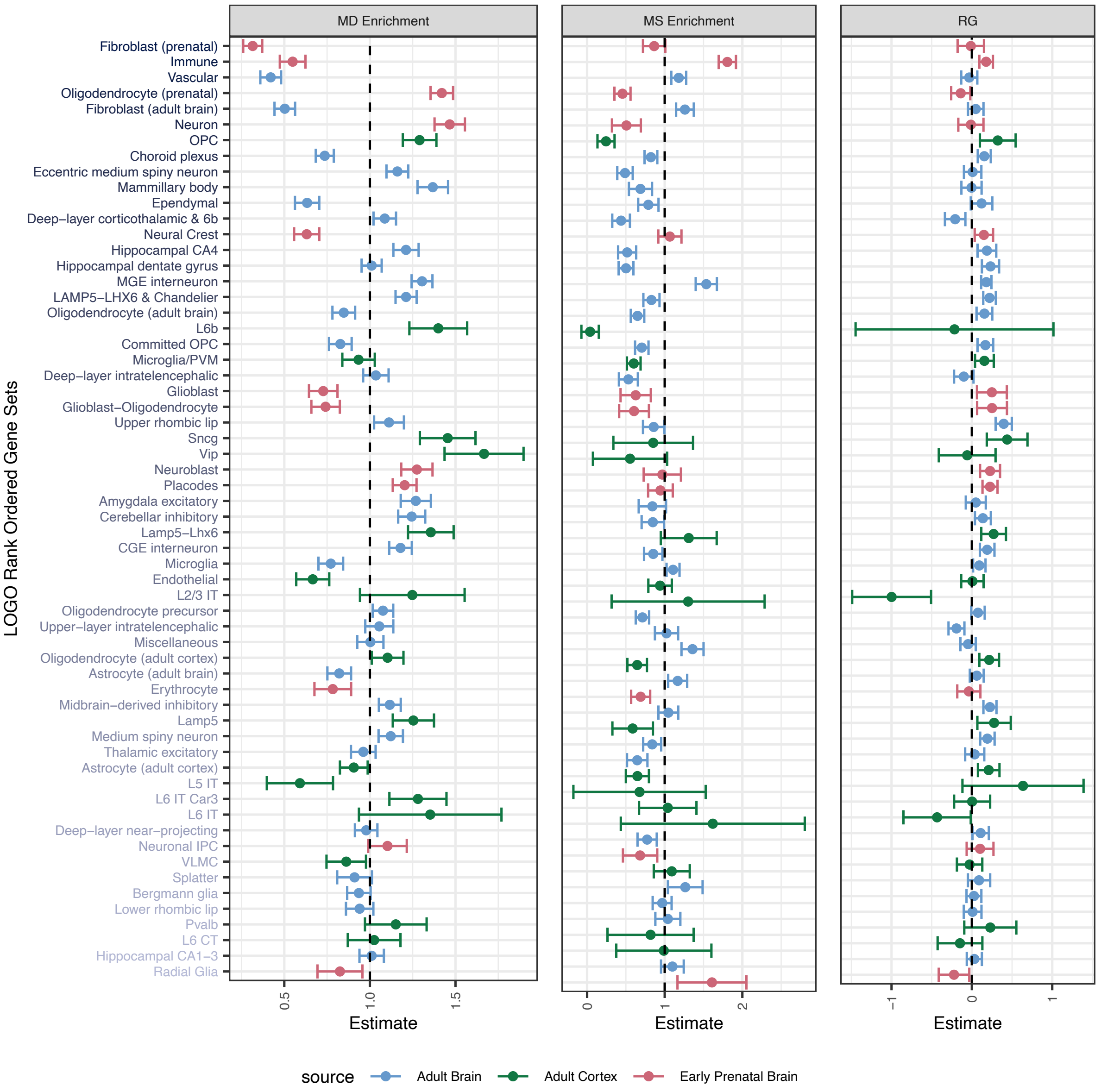


**Supplementary Figure 12. MD vs MS LoGO Rankings**. Depicted is the cell type gene sets on the y-axis ranked and colored based on the strength of the LoGO statistic. Each panel displays either the trait specific cell type enrichment or the local $r_g$ for a given cell type.

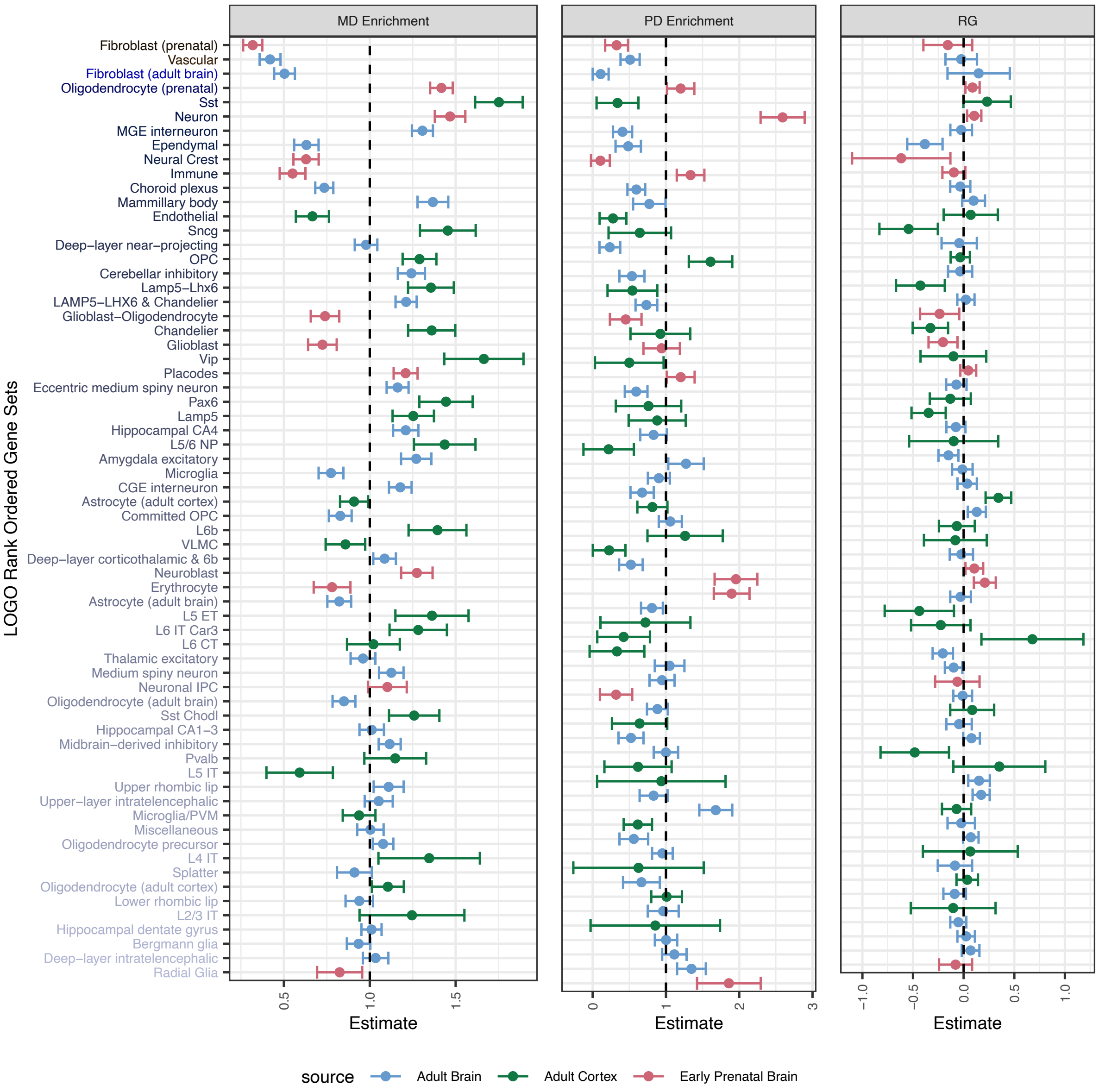


**Supplementary Figure 13. MD vs PD LoGO Rankings**. Depicted is the cell type gene sets on the y-axis ranked and colored based on the strength of the LoGO statistic. Each panel displays either the trait specific cell type enrichment or the local $r_g$ for a given cell type.

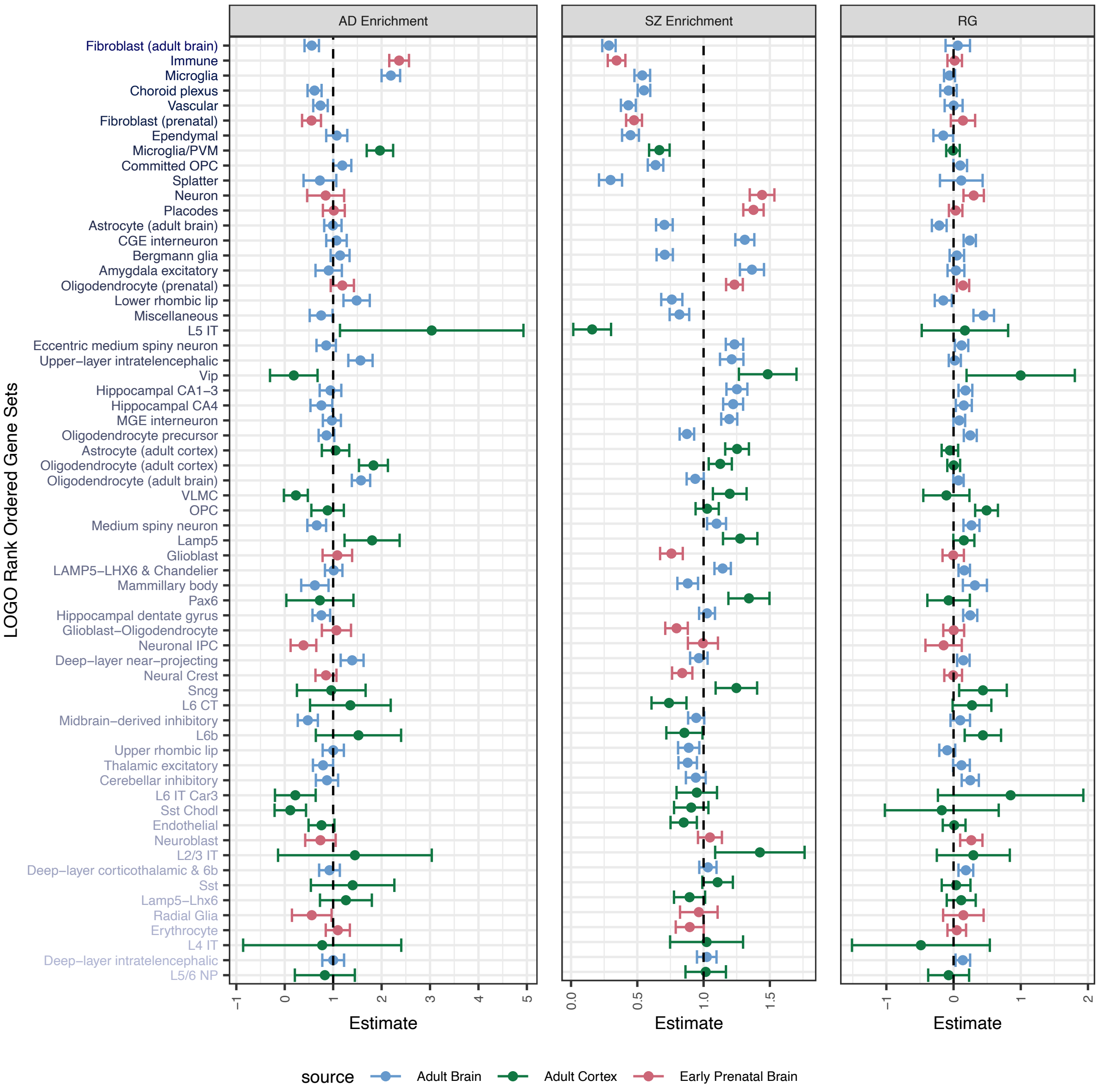


**Supplementary Figure 14. AD vs SZ LoGO Rankings**. Depicted is the cell type gene sets on the y-axis ranked and colored based on the strength of the LoGO statistic. Each panel displays either the trait specific cell type enrichment or the local $r_g$ for a given cell type.

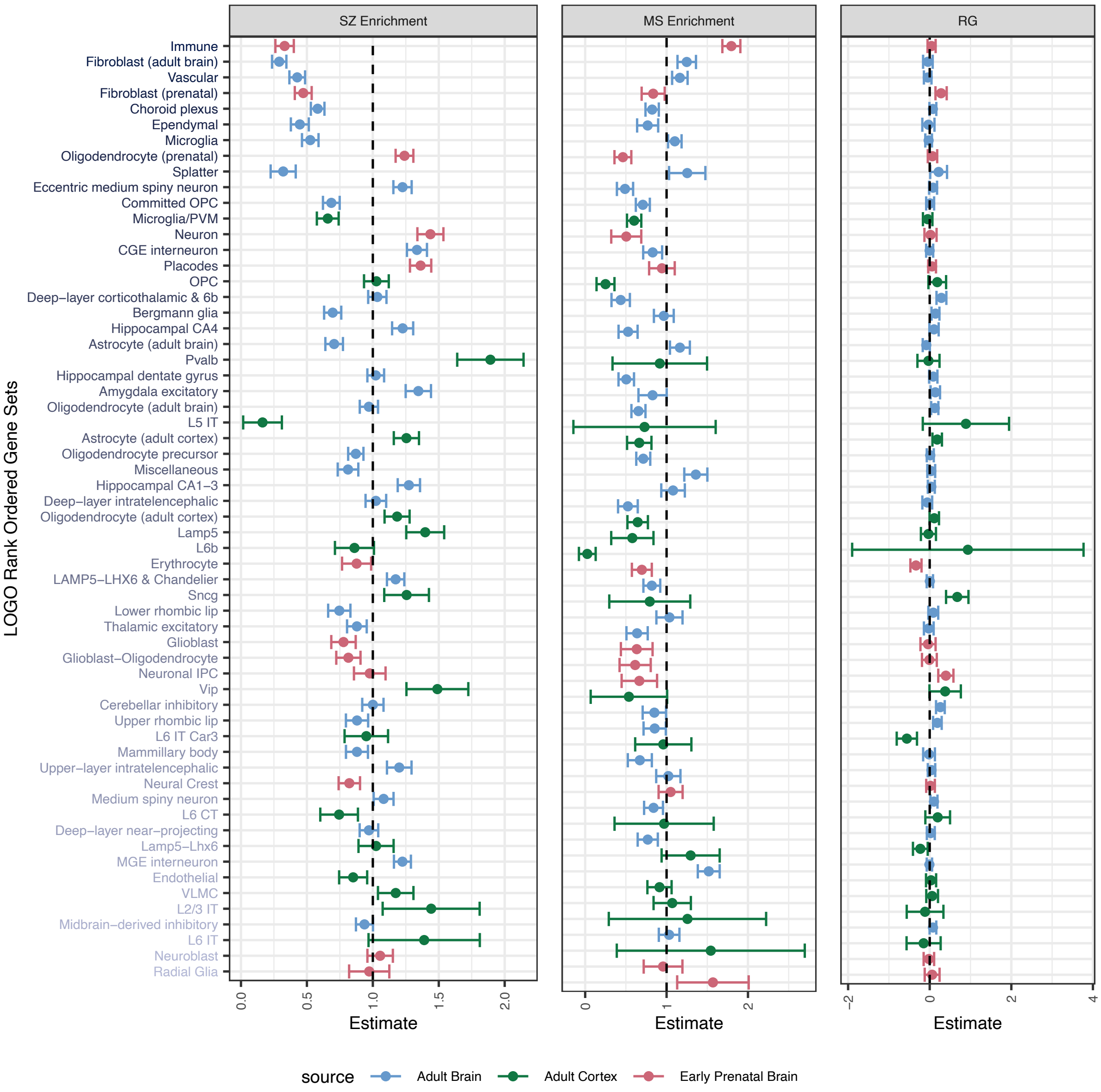


**Supplementary Figure 15. SZ vs MS LoGO Rankings**. Depicted is the cell type gene sets on the y-axis ranked and colored based on the strength of the LoGO statistic. Each panel displays either the trait specific cell type enrichment or the local $r_g$ for a given cell type.

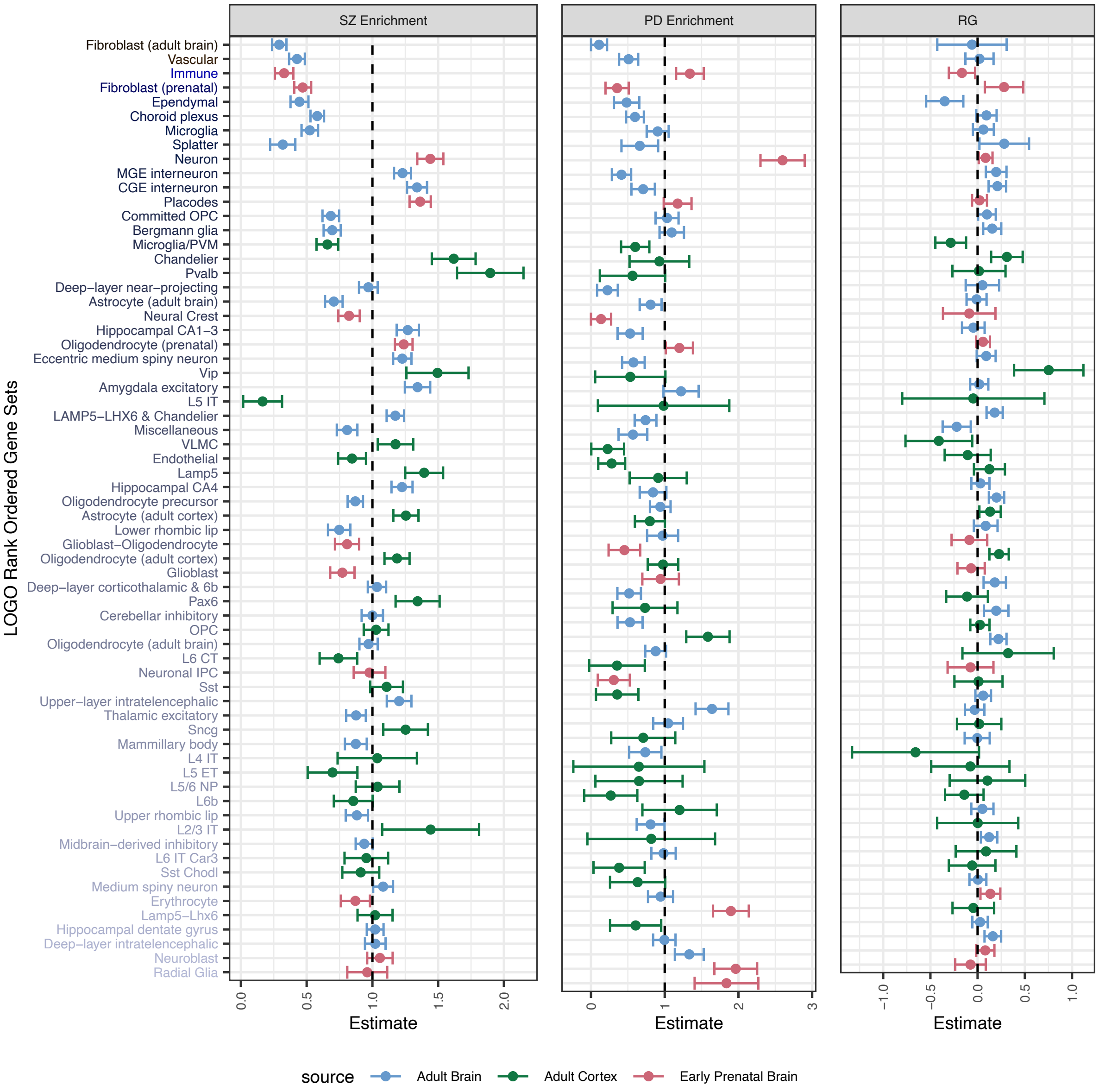


**Supplementary Figure 16. SZ vs PD LoGO Rankings**. Depicted is the cell type gene sets on the y-axis ranked and colored based on the strength of the LoGO statistic. Each panel displays either the trait specific cell type enrichment or the local $r_g$ for a given cell type.

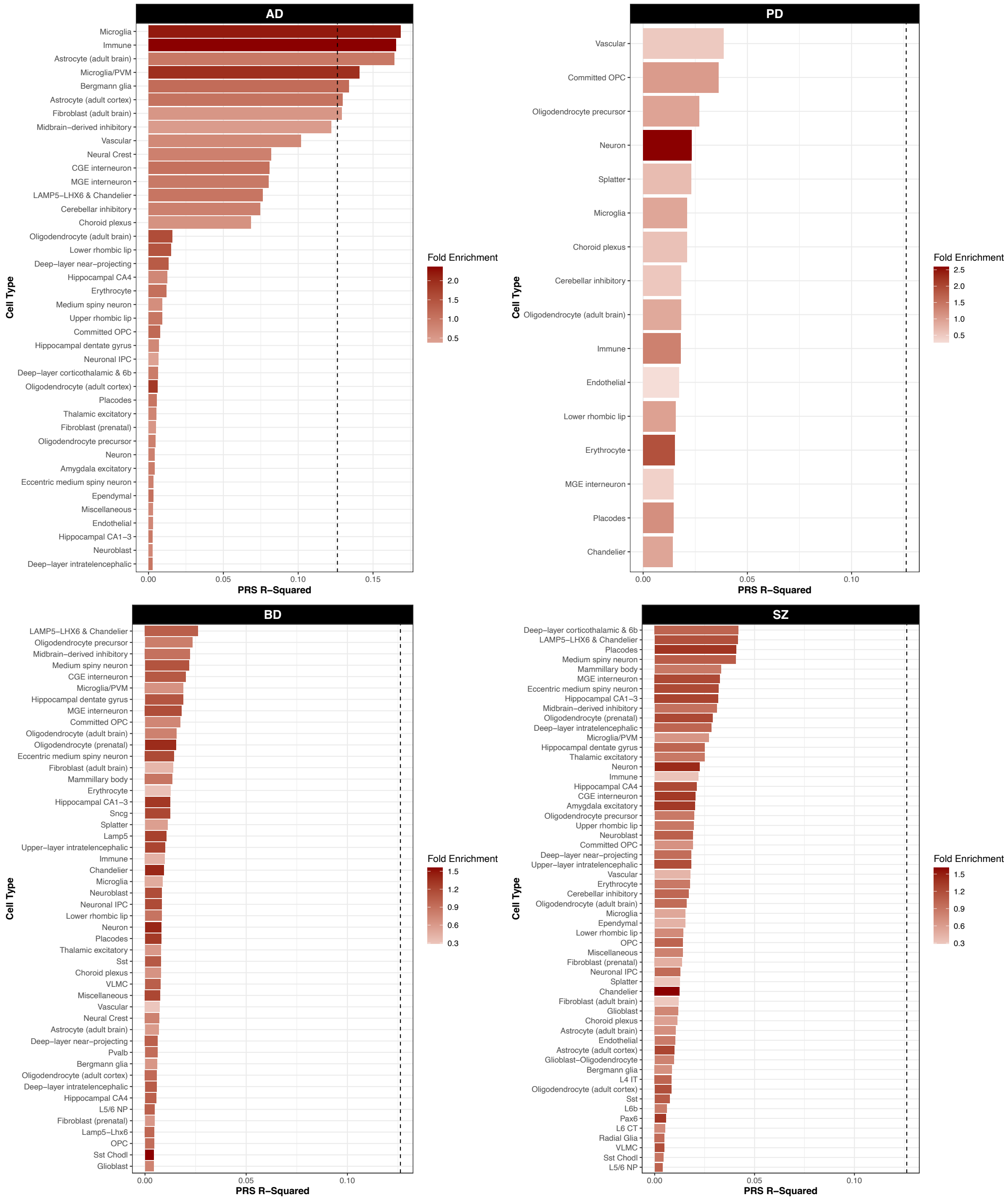


**Supplementary Figure 17. Univariate Cell Type Polygenic Risk Scores.** Variance explained by univariate cell type polygenic risk scores (PRS) with the intensity of the bar color represented by the strength of fold enrichment.

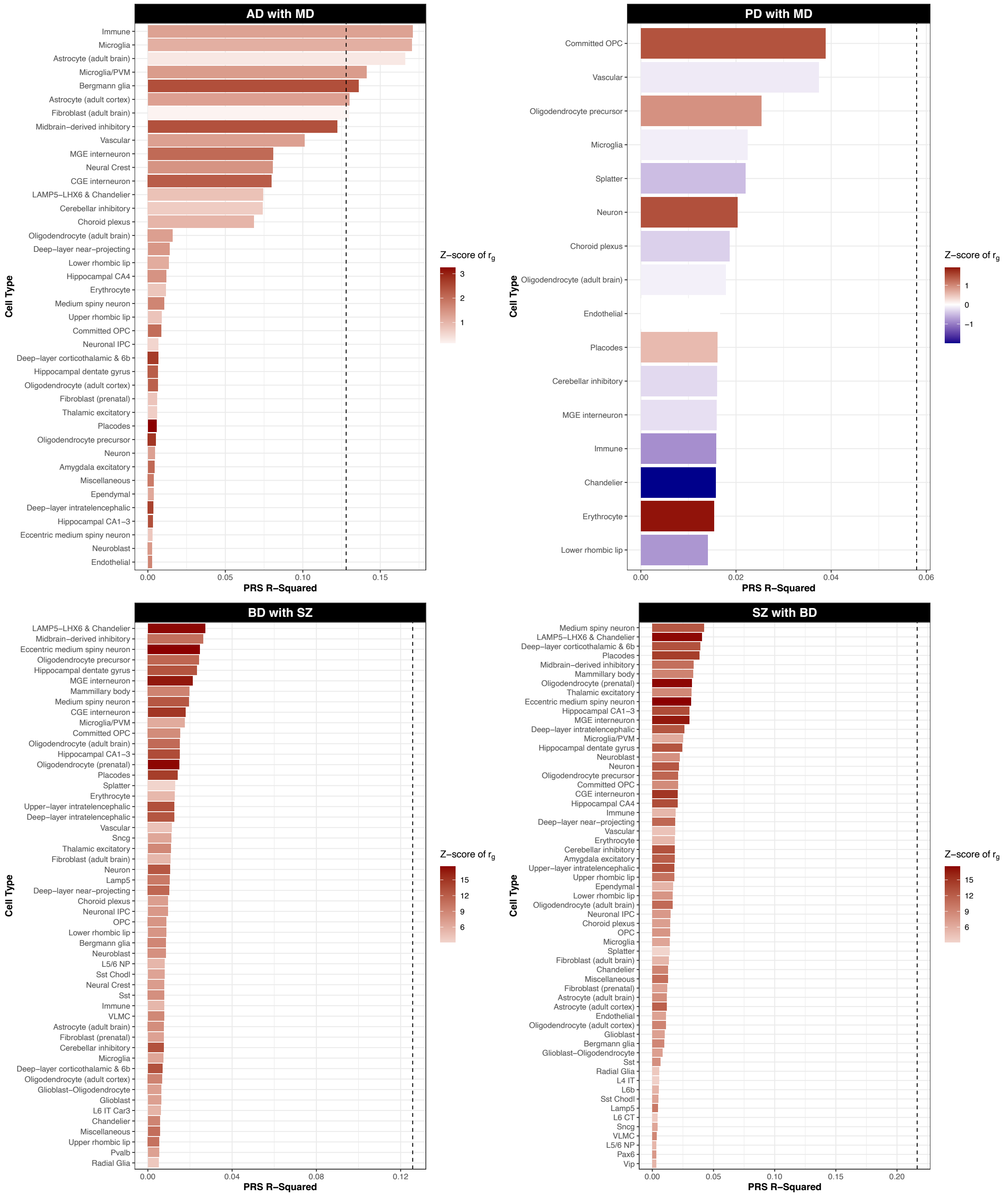


**Supplementary Figure 18. Bivariate Cell Type Polygenic Risk Scores with Local Genetic Correlations.** Variance explained by bivariate cell type PRS where the secondary trait is selected based on the average absolute value of the z-score for local $r_g$ across cell type gene sets. The intensity of the bar color represents the strength of the local $r_g$ between the two traits for the given cell type. The correlation between the PRS $R^2$ and the absolute value of the z-score for $r_g$ was as follows: AD with MD r=-0.23, p= 0.15; PD with MD r=-4.18e-4, p=1.00; BD with SZ r= 0.57, p= 1.05e-05; and SZ with BD r= 0.67, p= 7.25e-09.

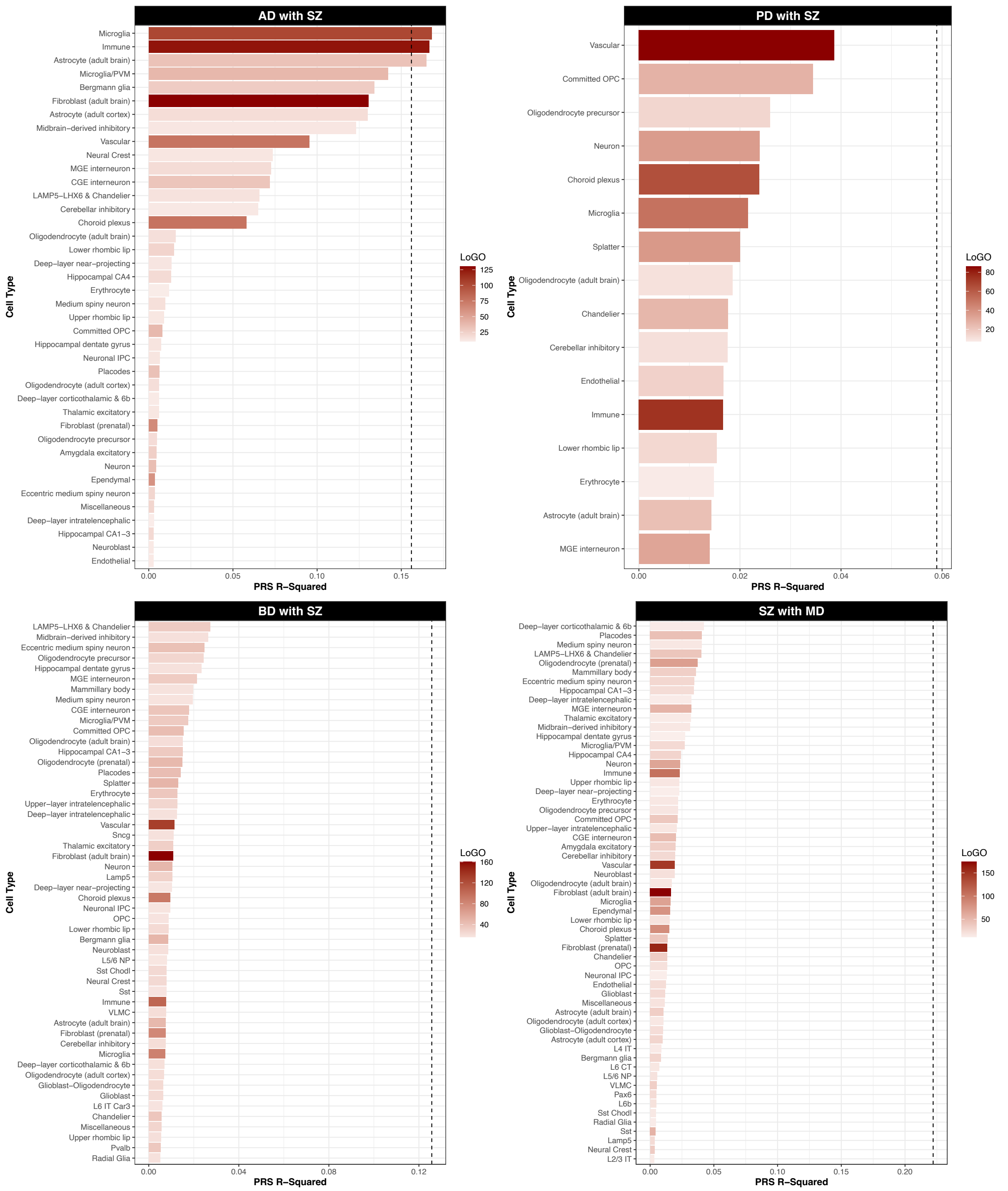


**Supplementary Figure 19. Bivariate Cell Type Polygenic Risk Scores with Local Genetic Omnibus (LoGO) Statistic**. Variance explained by bivariate cell type PRS where the secondary trait is selected based on the average LoGO statistic across cell type gene sets. The intensity of the bar color represents the strength of the LoGO statistic between the two traits for the given cell type. The correlation between the PRS $R^2$ and the LoGO statistic was as follows: AD with SZ r=0.54, p=3.21e-4; PD with SZ r=-0.49, p=0.05; BD with SZ r=-0.06, p=0.63; or SZ with MD r=0.02, p=0.88.
