## Supplementary Methods for "Bivariate GSA-MiXeR: A Novel Tool for Functional Genomic Analyses Implicates Diverse Neural Cell Types for Psychiatric and Neurodegenerative Disorders"

### Bivariate GSA MiXeR - Supplementary Note 1.

#### Contents


**1 Simple additive genetic model for complex traits**

1.1 Probability model for GWAS z-scores . . . . . . . . . . . . . . . . . . . . .

**2 Heritability models (univariate analysis)**

2.1 Null model with MAF-dependent architecture . . . . . . . . . . . . . . .

2.2 Baseline model with functional annotations . . . . . . . . . . . . . . . . . .

2.2.1 Fold enrichment of heritability . . . . . . . . . . . . . . . . . . . . .

2.2.2 Log-likelihood function with functional annotations . . . . . . .

2.3 Full model with genomic regions of interest . . . . . . . . . . . . . . . . . .

**3 Bivariate models**

3.1 Bivariate model with partitioned covariance . . . . . . . . . . . . . . .

3.2 Implementation details . . . . . . . . . . . . . . . . . . . . . . . . . . . . .

### 1 Simple additive genetic model for complex traits

Assuming a simple additive genetic model, a quantitative phenotype $y \in \mathbb{R}$ is modeled as a linear combination of genotype dosages $g_i$ with weights $\beta_i$:

$$y = \sum_{i=1}^{\bar{M}} g_i \beta_i + \epsilon_y \ ,$$

where the index $i$ runs over $\bar{M}$ genetic variants, weights $\beta_i \in \mathbb{R}$ are known as *additive effects of allele substitution*, and the $\epsilon_y \sim N(0, 1-h^2)$ term is a normally distributed residual, which reflects contributions from the environment and non-additive genetic effects. The quantitative phenotype $y$ is assumed to be centered and scaled, $E[y] = 0, \mathrm{Var}[y] = 1$, implying that the variance $\mathrm{Var}[\sum_i \beta_i g_i]$ is equal to each trait's SNP heritability, $h^2 \in [0,1]$. Genotype dosages $g_i$ are assumed to be centered but not scaled, i.e., $g_i \in \{0-2f_i, 1-2f_i, 2-2f_i\}$, where $f_i$ is allele frequency of the $i$-th variant.

Specific prior distribution on effect sizes $\beta_i$ encodes various genetic architectures of complex traits, for example $\beta_i \sim N(0, \sigma^2_\beta)$ assumption is widely known as *infinitesimal model*. Here $N(\mu, \sigma^2)$ denotes a random variable with Gaussian distribution with mean $\mu$ and variance $\sigma^2$, and we also let $N(x|\mu, \sigma^2) = \frac{1}{\sigma\sqrt{2\pi}} \exp(-\frac{(x-\mu)^2}{2\sigma^2})$ denote the probability density of a normal distribution at point $x$. Under infinitesimal model, $\sigma^2_\beta$ parameter is closely related to trait's heritability, $h^2 = \sum_i \sigma^2_\beta H_i$, where $H_i = 2f_i(1-f_i)$ is the heterozygosity of $i$-th variant. Indeed, by definition of heritability:

$$h^2 = \mathrm{Var}(\sum_{i=1}^{\bar{M}} g_i \beta_i)/\mathrm{Var}(y).$$

Then, as $g_i$ and $\beta_i$ are independent zero mean random variables,

$$h^2 = \sum_{i=1}^{\bar{M}} \mathrm{Var}(g_i)\mathrm{Var}(\beta_i)/\mathrm{Var}(y) = \sum_{i=1}^{\bar{M}} H_i \sigma^2_\beta$$

(as $\mathrm{Var}(g_i) = H_i$, $\mathrm{Var}(\beta_i) = \sigma^2_\beta$, and $\mathrm{Var}(y) = 1$).

Various extensions of this basic model are possible by modifying the prior distribution of effect sizes $\beta_i$, allowing the distribution of heritability to depend on SNP's allele frequency, functional annotations, genes or gene-sets, and we further extend this model to two traits, modeling partitioning genetic covariance across genomic regions of interest (ROI). We will describe these extensions in the following sections, and refer to them as *heritability models*, as they define how heritability is distributed across the genome.

#### 1.1 Probability model for GWAS z-scores

To infer numeric parameters defining the heritability model (e.g. $\sigma^2_\beta$ as in $\beta \sim N(0, \sigma^2_\beta)$ we use z-scores $(z_j)$ from GWAS summary statistics, where the index $j = 1, \ldots, M$

runs over GWAS tag SNPs (imputed or genotyped). Note that the set of GWAS tag SNPs must be a subset of the variants used in modeling additive genetic effects $\beta_i$, with $i = 1, \dots, \bar{M}$.

The following equations relate additive effects of allele substitution $\beta_i$ to $z_j$-scores from GWAS summary statistics.

*Let $\hat{\beta}'_j$ be GWAS estimate of the marginal effect size for $j$-th tag SNP, assessed via univariate linear regression, and $z_j$ be the corresponding z-score, $z_j = \hat{\beta}'_j / \hat{se}(\beta'_j)$. Then*

$$\begin{aligned} z_j &= \sqrt{N_j}\delta_j + \epsilon_j, \\ \delta_j &= \sum_{i=1}^{\bar{M}} a_{ij}\beta_i, \ \text{where } a_{ij} = \sqrt{H_i} r_{ij}, \\ \epsilon_j &\sim \mathcal{N}(0, \omega_0^2)\ , \end{aligned} \tag{1}$$

*where $N_j$ is the number of subjects with non-missing genotype information on $j$-th variant; $H_i = 2f_i(1 - f_i)$ is the heterozygosity of $i$-th variant; $r_{ij} = corr(\mathbf{v_i}, \mathbf{v_j})$ is an LD allele count correlation between genotype vectors $\mathbf{v_i}$, and $\mathbf{v_j}$ (two column-vectors running across individuals, containing dosages $g_i$ and $g_j$, of variants $i$ and $j$ respectively, across all individuals), parameter $\omega_0^2$ accounts for non-polygenic inflation in GWAS z-scores, and $\epsilon_j$ denotes a normally distributed residual.*

In a matrix form:

$$\mathbf{z} = \mathbf{N}^{1/2}\,\boldsymbol{R}\,\mathbf{H}^{1/2}\,\boldsymbol{\beta} + \boldsymbol{\epsilon}, \qquad \boldsymbol{\epsilon} \sim \mathcal{N}(\mathbf{0}, \omega_0^2 I_M), \tag{2}$$

where

$\mathbf{z} = (z_1, \dots, z_M)^\top$, vector of GWAS z-scores
$\mathbf{N}^{1/2} = \mathrm{diag}\big(\sqrt{N_1}, \dots, \sqrt{N_M}\big)$, $N_j$ is per-SNP effective sample size for tag SNP $j$,
$\mathbf{H}^{1/2} = \mathrm{diag}\big(\sqrt{H_1}, \dots, \sqrt{H_{\bar{M}}}\big)$, $H_i = 2f_i(1 - f_i)$ is heterozygosity of the $i$-th modeled variant,
$\mathbf{R} = (r_{ij})_{\substack{j=1,\dots,M \\ i=1,\dots,\bar{M}}}$ is an LD correlation matrix between tag SNPs (rows) and modeled variants (colu[illegible]
$\boldsymbol{\beta} = (\beta_1, \dots, \beta_{\bar{M}})^\top$,
$\boldsymbol{\epsilon} \sim \mathcal{N}(\mathbf{0}, \omega_0^2 I_M)$ is independent noise.

Note that the heterozygosity vector $\mathbf{H}$ has length $\bar{M}$, matching the modeled-variant index of the columns in $\mathbf{R}$. When the set of tag SNPs coincides with the set of modeled variants ($M = \bar{M}$), $\mathbf{R}$ reduces to the familiar symmetric LD correlation matrix.

**Proof**. In the absence of covariates, the least squares estimate $\hat{\beta}'_j$ can be expressed as

$$\hat{\beta}'_j = \frac{\mathbf{v_j^T y}}{\mathbf{v_j^T v_j}} = \beta_j + \sum_{i \neq j} \hat{\xi}_{ij}\beta_i + \frac{\mathbf{v_j^T e}}{\mathbf{v_j^T v_j}} = \sum_i \sqrt{\frac{H_i}{H_j}} r_{ij}\beta_i + \frac{\epsilon_j}{\sqrt{H_j N_j}}\ ,$$

where $\hat{\xi}_{ij} = \mathbf{v_i^T v_j} / \mathbf{v_j^T v_j} = \hat{\zeta}_{ij} / \hat{\zeta}_{jj}$, with $\hat{\zeta}_{ij} = \mathbf{v_i^T v_j}/N$ being an estimate of the covariance between $i$-th and $j$-th variants:

$$\hat{\zeta}_{ij} \simeq \sqrt{2f_i(1 - f_i)}\sqrt{2f_j(1 - f_j)}\, r_{ij}\ .$$

Here, the symbol "$\simeq$" denotes asymptotic equality as $n \to \infty$ . Then using $\hat{se}(\beta'_j) = 1/\sqrt{H_j N_j}$ result in Eq. (1). ■

### 2 Heritability models (univariate analysis)

This section describes *null model* (accounting for MAF-dependent architecture), *baseline model* (additionally accounting for functional annotations), and *full model* (additionally accounting for genomic regions of interest, such as genes or gene-sets).

#### 2.1 Null model with MAF-dependent architecture

The null model is defined by $\beta_i \sim N(0, \sigma^2_\beta H_i^S)$, where the $S$ parameter controls the effect size distribution with regards to allele frequency, similarly to LDAK and SumHer models. We note that effect sizes $\beta_i$ are defined w.r.t. centered but not scaled genotypes $g_i$, so that $\mathrm{Var}(g_i) = H_i = 2f_i(1-f_i)$, thus under our null model trait's SNP heritability $h^2 = \sum_{i=1}^{\bar{M}} \sigma^2_\beta H_i^{S+1}$. The original GCTA model assumes $S = -1$, so that each SNP contributes equally to heritability regardless of its allele frequency.

#### 2.2 Baseline model with functional annotations

Baseline model allows heritability model to depend on known functional annotations, with per-SNP variance $\sigma_i^2$ expressed via the following expression:

$$\sigma_i^2 = \big(\sigma^2_{A,0} + \sum_{p=1}^{N_a} [i \in A_p] \sigma^2_{A,p}\big) H_i^S \tag{3}$$

The index $p$ runs across functional annotation categories $\{A_1, A_2, \ldots, A_{N_a}\}$, while the parameter $\sigma^2_{A,p}$ represents the contribution of the $p$-th annotation category to the variance of $i$-th genetic variant, given that the $i$-th variant belongs to the $p$-th category, as specified in 3 via the indicator variable $[i \in A_p]$, using square brackets notation to transform a logical statement from true or false into 1 and 0, respectively. If genetic variant $i$ belongs to multiple annotation categories, the variance parameters of those annotations will be added together in an additive manner. To simplify notation we will introduce "base" category $A_0$ including all variants, and run index $p = 0, \ldots, N_a$:

$$\sigma_i^2 = \sum_{p=0}^{N_a} [i \in A_p] \sigma^2_{A,p} \tag{4}$$

In a matrix notation this is equivalent to $\boldsymbol{\beta} \sim \mathcal{N}(\mathbf{0}, \Sigma_\beta)$ where

$$\Sigma_\beta \;=\; \mathrm{diag}\big(U \boldsymbol{\sigma}^2_A\big),$$

$$U = (u_{ip}) \in \{0,1\}^{M\times(N_a+1)}, u_{ip} = 1 \text{ iff SNP } i \in A_p,$$
$$\boldsymbol{\sigma}^2_A = (\sigma^2_{A,0}, \sigma^2_{A,1}, \ldots, \sigma^2_{A,N_a})^\top,$$
$$\text{so that} \quad \sigma^2_i = \sum_{p=0}^{N_a} u_{ip}\, \sigma^2_{A,p},$$
$$\text{and therefore} \quad \Sigma_\beta = \text{diag}(\sigma^2_1, \ldots, \sigma^2_M), \quad \text{Cov}(\beta_i, \beta_j) = 0 \ (i \neq j).$$

##### 2.2.1 Fold enrichment of heritability

Assuming all parameters are known or estimated from GWAS summary statistics, one can compute heritability (and its fold enrichment) for any given set of SNPs $G \subset \{1, \ldots, \bar{M}\}$:

$$h^2(G) = \text{Var}(\sum_{i\ \in G} g_i\beta_i)/\text{Var}(y) = \sum_{i\in G} H_i\sigma^2_i.$$

The fold enrichment $f(G)$ of heritability is defined as a proportion of trait's SNP heritability in $G$ estimated from the baseline model divided by the expected proportion of trait's heritability in $G$ estimated under the null model:

$$f(G) = \frac{h^2_{base}(G)/h^2_{base}}{h^2_{null}(G)/h^2_{null}} = \frac{\big(\sum_{i\in G} H_i\sigma^2_i\big)/\big(\sum_{i=1}^{\bar{M}} H_i\sigma^2_i\big)}{\big(\sum_{i\in G} H_i^{S+1}\big)/\big(\sum_{i=1}^{\bar{M}} H_i^{S+1}\big)}.$$

More explicitly, this can be written as follows:

$$f(G) = \frac{\big(\sum_{i\in G}\sum_{p=0}^{N_a}[i \in A_p] H_i^{S+1}\sigma^2_{A,p}\big)/\big(\sum_{i=1}^{\bar{M}}\sum_{p=0}^{N_a}[i \in A_p] H_i^{S+1}\ \sigma^2_{A,p}\big)}{\big(\sum_{i\in G} H_i^{S+1}\big)/\big(\sum_{i=1}^{\bar{M}} H_i^{S+1}\big)}.$$

Note that in this case fold enrichment is computed for the basleine model w.r.t. the null model. Later we're interested in fold enrichment of the full model, which accounts for genomic regions of interest (e.g. genes or gene-sets), w.r.t baseline model.

##### 2.2.2 Log-likelihood function with functional annotations

The log-likelihood function of observing GWAS z-scores $\mathbf{z} = (z_j)$ given model parameters, $\theta = (\{\sigma^2_{A,p}\}, \omega_0)$, can be written as follows:

$$\begin{aligned}\log L(\mathbf{z}|\theta) &= \sum_j w_j \log\phi(z_j; 0, \omega_0^2 + N_j \sum_{i=1}^{\bar{M}} a^2_{ij}\sigma^2_i) = \\ &= \sum_j w_j \log\phi(z_j; 0, \omega_0^2 + N_j \sum_{p=0}^{N_a}\sum_{i=1}^{\bar{M}} a^2_{ij}[i \in A_p] H_i^S\sigma^2_{A,p}) = \qquad (5)\\ &= \sum_j w_j \log\phi(z_j; 0, \omega_0^2 + N_j \sum_{p=0}^{N_a} \ell_{jp}\sigma^2_{A,p}),\end{aligned}$$

where at the last step we've used expression for $a_{ij}$, and introduced heterozygosity-adjusted LD scores $\ell_{jp}$ for each functional category $p$ and tag SNP $j$:

$$\ell_{jp} = \sum_{i=1}^{\bar{M}} a_{ij}^2 [i \in A_p] H_i^S = \sum_{i \in A_p} H_i^{S+1} r_{ij}^2. \tag{6}$$

Weights $w_j$ are introduced to avoid over-counting contribution from large LD blocks, and $w_j$ can be computed from inverse LD weighting (default), or as probabilities of selecting $j$-th SNP in random pruning procedure; $\phi(z; 0, s^2) = \frac{1}{\sqrt{2\pi s}} e^{-z^2/2s^2}$ stands for the probability density function of centered normal distribution, $N_j$ is GWAS sample size for SNP $j$.

Under fixed value of $S$ parameter, equation (5), $E[z_j^2] = \omega_0^2 + N_j \sum_{p=0}^{N_a} \ell_{jp} \sigma_{A,p}^2$, is closely related to stratified LDSC regression model, with regression parameters $\vec{x} = \sigma_{A,p}^2$ and intercept $\omega_0^2$.

#### 2.3 Full model with genomic regions of interest

Full model extends baseline model (3) by additional multiplicative term which accounts for genomic regions of interest (genomic ROIs), such as genes or gene-sets:

$$\sigma_i^2 = \left(\sigma_{A,0}^2 + \sum_{p=1}^{N_a} [i \in A_p] \sigma_{A,p}^2\right) \left(\sigma_{G,0}^2 + \sum_{q=1}^{N_g} [i \in G_q] \sigma_{G,q}^2\right) H_i^S, \tag{7}$$

where index $q$ runs across genomic ROIs $\{G_1, G_2, \ldots, G_{Ng}\}$, while the parameter $\sigma_{G,q}^2$ represents the contribution of $q$-th genomic ROI. $\sigma_{G,0}^2$ parameter ensures non-zero variance $\sigma_i^2$ for genetic variants $G_0$ that do not belong to any genomic ROI.

We note that the way of extending baseline model into full model is mathematically quite similar to the way baseline model extended the null model:

$$\begin{aligned} \sigma_{full,i}^2 &= \sigma_{base,i}^2 \sum_{q=0}^{N_g} [i \in G_q] \sigma_{G,q}^2, \text{ and} \\ \sigma_{base,i}^2 &= \sigma_{null,i}^2 \sum_{p=0}^{N_a} [i \in A_p] \sigma_{A,p}^2, \end{aligned} \tag{8}$$

thus the same formulas for log-likelihood function and software modules can be used for estimating log-likelihood, fitting parameters of the full model while keeping baseline model parameters fixed, and for estimating fold enrichments $f(G) = \frac{h_{full}^2(G)/h_{full}^2}{h_{base}^2(G)/h_{base}^2}$ for genomic ROIs $G$, between the full model and the baseline models.

### 3 Bivariate models

Simple additive genetic model for two quantitative traits $y_1, y_2$ can be written as follows:

$$\begin{aligned} y_1 &= \sum_{i=1}^{\bar{M}} g_i \beta_{1i} + \epsilon_1, \\ y_2 &= \sum_{i=1}^{\bar{M}} g_i \beta_{2i} + \epsilon_2, \end{aligned} \tag{9}$$

where $g_i$ is an additively coded number of reference alleles for $i$-th variant, $\beta_{1i}$ ($\beta_{2i}$) is the effect of $i$-th variant on, respectively, trait $y_1$ ($y_2$), and $\epsilon_1$ ($\epsilon_2$) is the residual component. Here we choose that random variables $y_1$, $\beta_{1j}$ and $\epsilon_1$ are centered and scaled so that $E(y_1) = 0$ and $\mathrm{Var}(y_1) = 1$, and likewise for the second trait. We allow for dependencies between genetic effects for same variant $(\beta_{1j}, \beta_{2j})$, and also allow for dependency between the residual $(\epsilon_1, \epsilon_2)$. Apart from these dependencies, we model genetic effects $\beta_{1j}$ and $\beta_{2k}$ ($j \neq k$) independently from each other, and independently from the environmental terms $\epsilon_1, \epsilon_2$.

For each trait, its *heritability* is defined as the variance of the genetic component:

$$h_1^2 = \frac{\mathrm{Var}(\sum_{j=1}^{\bar{M}} g_j \beta_{1j})}{\mathrm{Var}(y)} = \sum_{j=1}^{\bar{M}} H_j \mathrm{Var}(\beta_{1j}), \tag{10}$$

where $H_j = 2p_j(1-p_j)$ is heterozygosity of $j$-th variant. Likewise, for the second trait $h_2^2 = \sum_{j=1}^{\bar{M}} H_j \mathrm{Var}(\beta_{2j})$. *Genetic correlation* between the traits is defined as the covariance of the genetic components, normalized by $\sqrt{h_1^2 h_2^2}$:

$$\rho_g = \frac{\mathrm{Cov}(\sum_{j=1}^{\bar{M}} g_j \beta_{1j}, \sum_{j=1}^{\bar{M}} g_j \beta_{2j})}{\sqrt{h_1^2 h_2^2}} = \frac{1}{\sqrt{h_1^2 h_2^2}} \sum_{j=1}^{\bar{M}} H_j \mathrm{Cov}(\beta_{1j}, \beta_{2j}). \tag{11}$$

Allowing each trait to have its own heritability model, $\beta_{i1} \sim N(0, \sigma_{1i}^2)$ and $\beta_{i2} \sim N(0, \sigma_{2i}^2)$, with $\sigma_{1i}^2$ and $\sigma_{2i}^2$ defined based on null, baseline or full model as in previous sections. We introduce global genetic correlation parameter, $\rho$, and derive the following bivariate distribution on GWAS z-scores:

$$\begin{aligned} &(\beta_{1i}, \beta_{2i}) \sim \mathcal{N}(0, \mathbf{\Sigma}_i), \\ &\mathbf{\Sigma}_i = \begin{bmatrix} \sigma_{1i}^2 & \rho\sigma_{1i}\sigma_{2i} \\ \rho\sigma_{1i}\sigma_{2i} & \sigma_{2i}^2 \end{bmatrix}, \\ &(z_{1j}, z_{2j}) \sim (\delta_{1j}, \delta_{2j}) + \mathcal{N}\Big(0, \mathbf{\Omega}_0\Big), \\ &\delta_{\cdot j} = \sqrt{N_{\cdot j}} \sum_i \sqrt{H_i} r_{ij} \beta_{\cdot i}, \\ &\mathbf{\Omega}_0 = \begin{bmatrix} \omega_{01}^2 & \rho_0\omega_{01}\omega_{02} \\ \rho_0\omega_{01}\omega_{02} & \omega_{02}^2 \end{bmatrix}. \end{aligned} \tag{12}$$

#### 3.1 Bivariate model with partitioned covariance

The following model allows genetic correlation to vary across genomic regions of interest. Assuming no overlap between genomic regions, we define region-specific genetic correlation $\rho_q$ defining genetic covariance between the traits $\mathrm{Cov}(\beta_{1i}, \beta_{2i})$ as follows:

$$\begin{aligned}
&(\beta_{1i}, \beta_{2i}) \sim \mathcal{N}\Big(0, \sum_{q=0}^{N_g} [i \in G_q] \mathbf{\Sigma}_q\Big), \\
&\mathbf{\Sigma}_q = \begin{bmatrix} \sigma_{1i}^2 & \rho_q \sigma_{1i} \sigma_{2i} \\ \rho_q \sigma_{1i} \sigma_{2i} & \sigma_{2i}^2 \end{bmatrix}, \\
&(z_{1j}, z_{2j}) \sim (\sqrt{N_{1j}} \delta_{1j}, \sqrt{N_{2j}} \delta_{2j}) + \mathcal{N}\Big(0, \mathbf{\Omega}_0\Big), \\
&\delta_{1j} = \sum_i a_{ij} \beta_{1i}, \quad \delta_{2j} = \sum_i a_{ij} \beta_{2i}, \\
&\mathbf{\Omega}_0 = \begin{bmatrix} \omega_{01}^2 & \rho_0 \omega_{01} \omega_{02} \\ \rho_0 \omega_{01} \omega_{02} & \omega_{02}^2 \end{bmatrix}.
\end{aligned} \tag{13}$$

Full expression for log-likelihood is as follows:

$$\log L = \sum_j w_j \log \Phi(z_{1j}, z_{2j}; 0, \mathbf{\Omega}_0 + \sum_{q=0}^{N_g} \sum_{i=1}^{\bar{M}} a_{ij}^2 [i \in G_q] N_j^\top \mathbf{\Sigma}_q N_j), \tag{14}$$

where $\Phi(z_1, z_2; 0, \mathbf{\Sigma})$ is probability density of centered bivariate normal distribution with variance-covariance matrix $\mathbf{\Sigma}$, and $N_j = (\sqrt{N_{1j}}, \sqrt{N_{2j}})^T$ is a vector of length 2 with square roots of sample size of the $j$-th SNP.

*LoGO* statistic is defined as the difference between log-likelihood under full versus baseline models:

$$LoGO = \log L_{full} - \log L_{base},$$

where $\log L_{full}$ is bivariate log-likelihood function (14) modeling specific contributions of the genomic ROIs to heritability model of both traits, and their ROI-specific genetic correlations, while $\log L_{base}$ model is agnostic to genomic regions of interest. Both of these models (full and baseline) control for differential enrichment across functional annotations and MAF-dependent genetic architectures.

#### 3.2 Implementation details

Optimization procedure starts by fitting univariate parameters of the baseline and full models, separately for each trait. Then, parameters of the baseline and full bivariate models are optimized, while keeping all parameters of the univariate models fixed. At each step, method of moments (MoM) is used to initialize starting point for direct log-likelihood optimization (MLE), with non-negative least squares solution used in the univariate MoM, and ordinary least squares (OLS) used in bivariate MoM. MLE is implemented using PyTorch library, with `torch.autograd` feature to derive gradients for Adam first-order gradient-based optimization. For the full models, the intercept parameters $\omega_0$ and $\rho_0$ were estimated using their baseline model values as initial approximation. The full model is optimized for a subset of genomic regions of interest where MoM estimates were within parameter boundaries ($\sigma_g^2 > 0$, $|\rho_g| < 1$); for genomic regions with out-of-bound estimates the results are reported as missing values. In MLE estimation variance parameters are optimized in `log` space, and correlations parameters are optimized in `atanh` space, leading to unbounded optimization while keeping variance estimates positive and correlations constrained to $(-1, 1)$ range.

**Algorithm 1:** MLE workflow for baseline and full models

**Data:** GWAS z-scores $(z_{1j}, z_{2j})$, raw LD matrix $r_{ij}$, heterozygosities $H_i$, annotations $A_p$, genomic ROIs $G_q$, weights $w_j$, parameter $S$
**Result:** MLE estimates for baseline and full univariate/bivariate models

```
/* LD-score construction */
```
**foreach** *tag SNP j, annotation p* **do**
  $\ell_{jp} \leftarrow \sum_{i \in A_p} H_i^{S+1} r_{ij}^2$;
**end**

```
/* Univariate fitting per trait */
```
**foreach** *trait* $t \in \{1, 2\}$ **do**
  Fit baseline parameters $(\sigma^2_{A,p}, \omega_0)$ with MoM (NNLS on $\ell_{jp}$);
  **foreach** *tag SNP j, genomic ROI q* **do**
    $\ell_{jq} \leftarrow \sum_{i \in G_q} \sigma^2_{base,i} r_{ij}^2$;
  **end**
  Fit full MoM parameters (NNLS on $\ell_{jq}$);
  Optimize full model MLE $(\sigma^2_{G,q}, \omega_0)$ using Adam;
**end**

```
/* Bivariate fitting */
```
Fit baseline bivariate parameters $(\rho, \rho_0)$ with MoM (OLS on cross-trait $\ell_j$);
Fit full bivariate parameters $(\rho_q, \rho_0)$ with MoM (OLS on cross-trait $\ell_{jq}$);
Select ROIs with in-bounds MoM estimates (both traits $\sigma^2_{G,q} > 0$; $|\rho_g| < 1$);
Optimize full bivariate MLE $(\rho_q, \rho_0)$ with fixed univariate params;
Compute $LoGO = \log L_{full} - \log L_{base}$ (full vs baseline bivariate MLE);

```
/* Fold enrichment */
```
**foreach** *genomic ROI* $G_q$ **do**
  Compute $h^2_{\text{full}}(G_q)$ and $h^2_{\text{base}}(G_q)$ from fitted $\sigma^2_{A,p}$, $\sigma^2_{G,q}$;
  Evaluate $f(G_q) = \frac{h^2_{\text{full}}(G_q)/h^2_{\text{full}}}{h^2_{\text{base}}(G_q)/h^2_{\text{base}}}$;
**end**

```
/* Uncertainty */
```
Compute Hessian via `torch.autograd.functional.hessian`;
Invert negative Hessian for SEs; if not p.d., fall back to marginal errors;
Sample parameter values to propagate SEs into derived quantities, e.g. $f(G_q)$;

The log-likelihood $\log L(\theta|\mathbf{z})$ is also used to define standard errors (SEs) on parameters estimates via the observed Fisher information matrix, i.e. the negative of the Hessian matrix $\Delta\Delta^T \log L|_{\theta^*}$ of second partial derivatives of the log-likelihood function w.r.t. model parameters $\theta$ evaluated at the maximum likelihood estimates $\theta^*$:

$$\theta \sim N(\theta^*, [-\Delta\Delta^T \log L|_{\theta^*}]^{-1}).$$

The Hessian matrix is computed using `torch.autograd.functional.hessian` function. For compound functions of multiple parameters, such as fold enrichment of heritability in a genomic region, we sample N=100 realizations of the parameters vector from the above posterior distribution, calculate the compound function for each realization of the parameter vector, and report the standard deviations across the realizations. If the Hessian matrix was not positive definite, we use marginal errors of fitted parameters.

To speed up computation of bivariate normal zero-mean density one may take advantage of closed-form expression for the matrix inverse. This closed-form expression also allowed to use PyTorch library for gradient and Hessian estimation. Let variance matrix be $\mathbf{\Sigma} = \begin{bmatrix} a & b \\ b & c \end{bmatrix}$, then the precision matrix $\mathbf{\Sigma}^{-1} = \frac{1}{ac-b^2} \begin{bmatrix} c & -b \\ -b & a \end{bmatrix}$, and bivariate normal density function

$$\Phi(z_1, z_2; 0, \begin{bmatrix} a & b \\ b & c \end{bmatrix}) = \frac{1}{2\pi\sqrt{ac-b^2}} \exp\Big( -\tfrac{1}{2} \frac{cz_1^2 + az_2^2 - 2bz_1z_2}{ac-b^2} \Big). \tag{15}$$
